# Repeated introductions and intensive community transmission fueled a mumps virus outbreak in Washington State

**DOI:** 10.1101/2020.10.19.20215442

**Authors:** Louise H. Moncla, Allison Black, Chas DeBolt, Misty Lang, Nicholas R. Graff, Ailyn C. Pérez-Osorio, Nicola F. Müller, Dirk Haselow, Scott Lindquist, Trevor Bedford

## Abstract

In 2016/2017, Washington State experienced a mumps outbreak despite high childhood vaccination rates, with cases more frequently detected among school-aged children and members of the Marshallese community. We sequenced 166 mumps virus genomes collected during outbreaks in Washington and other US states, and apply phylodynamic approaches to trace mumps introductions and transmission within Washington. We uncover that mumps was introduced into Washington at least 13 times, primarily from Arkansas, sparking multiple co-circulating transmission chains. Neither vaccination status nor age were strong determinants of transmission. Instead, the outbreak in Washington was overwhelmingly sustained by transmission within the Marshallese community. Our findings underscore the utility of genomic data to clarify epidemiologic factors driving transmission, and pinpoint contact networks as critical determinants of mumps transmission. These results imply that contact structures and historic disparities may leave populations at increased risk for respiratory virus disease even when a vaccine is effective and widely used.

## Introduction

In 2016 and 2017, mumps virus swept the United States in the country’s largest outbreak since the pre-vaccine era (CDCMMWR, 2019). Washington State was heavily affected, reporting 889 confirmed and probable cases. Longitudinal studies (Davidkin et al., 2008), epidemiologic outbreak investigations (Cardemil et al., 2017), and epidemic models (Lewnard and Grad, 2018) suggest that mumps vaccine-induced immunity wanes over 13-30 years, consistent with the preponderance of young adult cases in recent outbreaks. Like with other recent mumps outbreaks, most Washington cases in 2016/17 were vaccinated. Unusually though, incidence was highest among children aged 10-18 years, younger than expected given waning immunity. The outbreak was also peculiar in that approximately 52% of the total cases were Marshallese, an ethnic community that comprises ∼0.3% of Washington’s population. These same phenomena were also observed in Arkansas. Of the 2,954 confirmed and probable Arkansas cases, 57% were Marshallese, and 57% of cases were children aged 5-17 (Fields et al., 2019). Amongst school-aged children in Arkansas and Washington, >90% had previously received 2 doses of MMR vaccine (Fields et al., 2019). The high proportion of vaccinated cases, younger-than-expected age at infection, disproportionate impact on the Marshallese community, and epidemiologic link to Arkansas suggest that factors beyond waning immunity are necessary to explain mumps transmission during this outbreak in Washington.

Between 1947 and 1986, the United States occupied the Republic of Marshall Islands and detonated the equivalent of >7000 Hiroshima size nuclear bombs as part of its nuclear testing program (Barker, 2012). The effects were devastating, precipitating widespread environmental destruction, nuclear contamination, and dire health consequences (Hallgren et al., 2015; Niedenthal, 1997; Palafox et al., 2007; Simon, 1997; Takahashi et al., 1997). Marshallese individuals inhabiting the targeted atolls were forcibly moved to other islands, and many were exposed to nuclear fallout (Abella et al., 2019) that persists on the Islands today (Bordner et al., 2016). Significant concern remains within the community regarding long-term health impacts of nuclear exposure and its potential impacts on immune function. Marshallese individuals living on and off the Islands experience significant health disparities including a higher burden from infectious diseases and chronic health conditions (Adams et al., 1986; Wong et al., 1979; Yamada et al., 2004). Compounding these disparities, from 1996 to 2020 (Hirono, 2019), Marshallese individuals were specifically excluded from Medicaid eligibility despite legal residency in the US permitted under the Compact of Free Association (COFA) Treaty. As a result, many US-residing Marshallese are uninsured, with poor access to healthcare (McElfish et al., 2015). Marshallese households also tend to be larger on average (Harris and Jones, 2005; US Census Bureau, n.d.), potentially increasing risk for respiratory virus exposure.

Sampling bias is a persistent challenge for elucidating source-sink dynamics during viral outbreaks. Here, we formulate a set of genomic epidemiological approaches that are robust to genomic sampling to investigate patterns of mumps transmission in Washington. We sequenced 110 mumps viral genomes obtained from specimens collected from laboratory-confirmed mumps cases in Washington State and another 56 from other US states collected between 2006 and 2018. We employ a novel application of phylogeographic methods to detailed epidemiologic data on age, vaccination status, and community membership, and develop a new statistic for quantifying transmission in the tree. By combining these phylodynamic approaches with community health advocate interviews that contextualize our results, we provide a framework for investigating viral transmission dynamics that is sensitive to community health priorities and readily applicable to other viral pathogens.

## Results

### Outbreak characteristics and dataset composition

We generated genome sequences for 110 PCR-positive mumps samples collected throughout Washington State during 2016/2017, and 56 samples collected in Wisconsin, Ohio, Missouri, Alabama, and North Carolina between 2006 and 2018 (**Supplemental Table 1**). The Washington State outbreak began in October 2016, and peaked in winter of 2017, culminating in 889 confirmed and probable cases across Washington (**Fig. 1**). Individuals aged <1 to 64 years were affected, but the highest rate of infection occurred in children aged 10-14 (44.9 cases per 100,000) and 15-19 (47.0 per 100,000) (**Supplemental Table 2**). Among individuals aged 5-19 years old, 91% were considered up-to-date on mumps vaccine. Adults in the age group most likely to be parents of school aged children (20-39 years old) were infected at a rate of only 12.9 cases per 100,000, but comprised a significant proportion (29%) of total cases (**Supplemental Table 2**). While Marshallese individuals comprise only ∼0.3% of Washington’s total population, they accounted for 52% of reported mumps cases (**Supplemental Table 3**). Among Marshallese individuals aged 5-19, 93% were up-to-date on vaccination, suggesting that this over-representation is not attributable to poor vaccine coverage.

**Figure 1:**
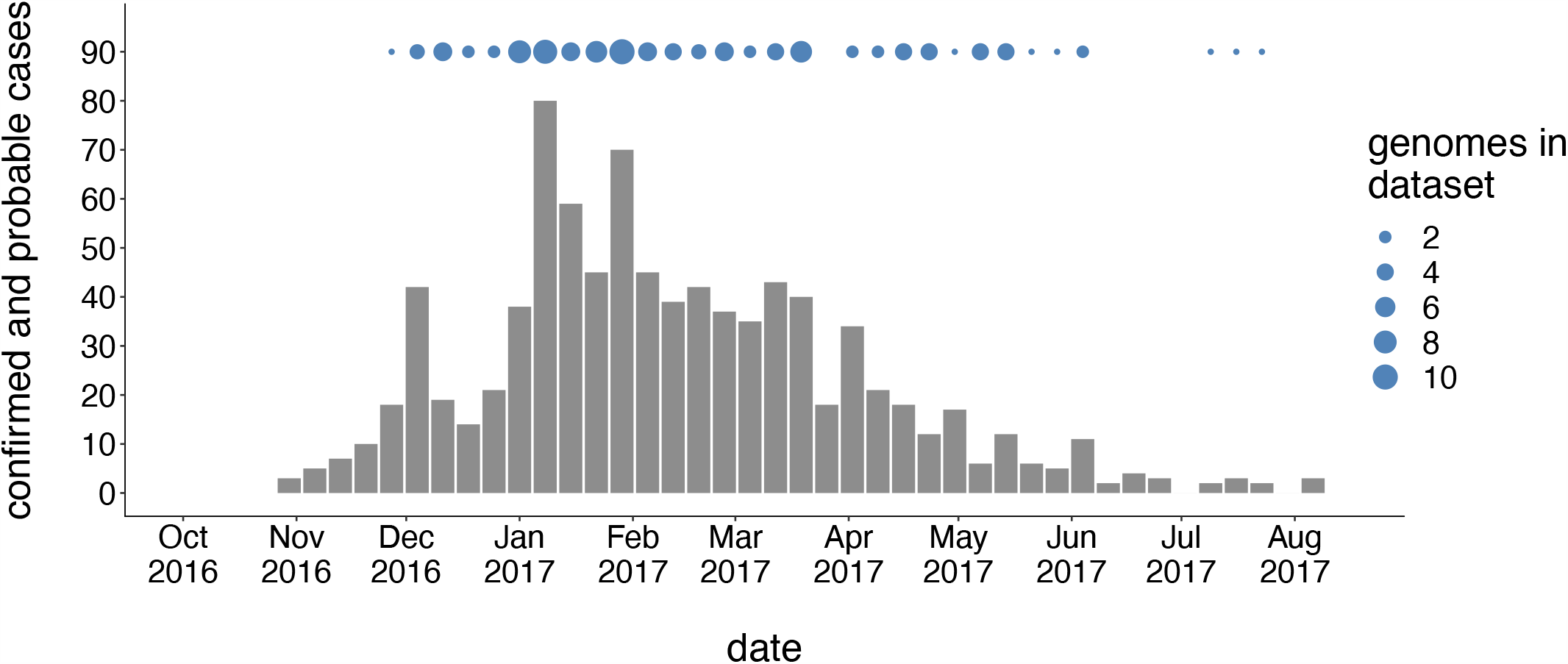
Washington case counts and genomic sampling. The first mumps case in Washington was reported on October 30, 2016, and case counts peaked in winter of 2017. The x-axis represents the date of the corresponding epidemiologic week, and the y-axis represents the number of confirmed and probable cases for that week. Blue dots above the case count plot represent the number of Washington genome sequences in our dataset sampled in that week.

### Outbreaks across North America are related

We combined our sequence data with publicly available full genome sequences sampled from North America between 2006 and 2018, and built a time-resolved phylogeny, inferring migration history among 26 US states and Canadian provinces (**Fig. 2, Supplemental Figure 1**). Sequences from samples collected between 2006 and 2014 clustered with other North American mumps viruses sampled from the same times. Except for 2 sequences (one from Wisconsin in 2006, genotype A, and one from Washington in 2017, genotype K, both excluded from **Fig. 2**), all samples in our dataset were genotype G viruses. Ten Washington sequences were highly divergent from other North American genotype G viruses, with a time to the most recent common ancestor (TMRCA) of ∼22 years (**Fig. 2**). The remaining Washington sequences nest within the diversity of other North American viruses, and descend from the same mumps lineage that has circulated in North America since 2006 (**Fig. 2**). We observe substantial geographic mixing along the tree. While viruses from Massachusetts (dark green tips and branches) seeded outbreaks in the Northeast and Midwest, we also infer transmission from Massachusetts to Texas, Louisiana, Alabama, and British Columbia. Despite the close geographic proximity between British Columbia and Washington, most British Columbia sequences form a distinct cluster on a long branch (**Fig. 2**), suggesting seeding from an unsampled location. Although viruses from Washington are scattered throughout the phylogeny, most cluster within a clade of viruses sampled in Arkansas (**Fig. 2**).

**Figure 2:**
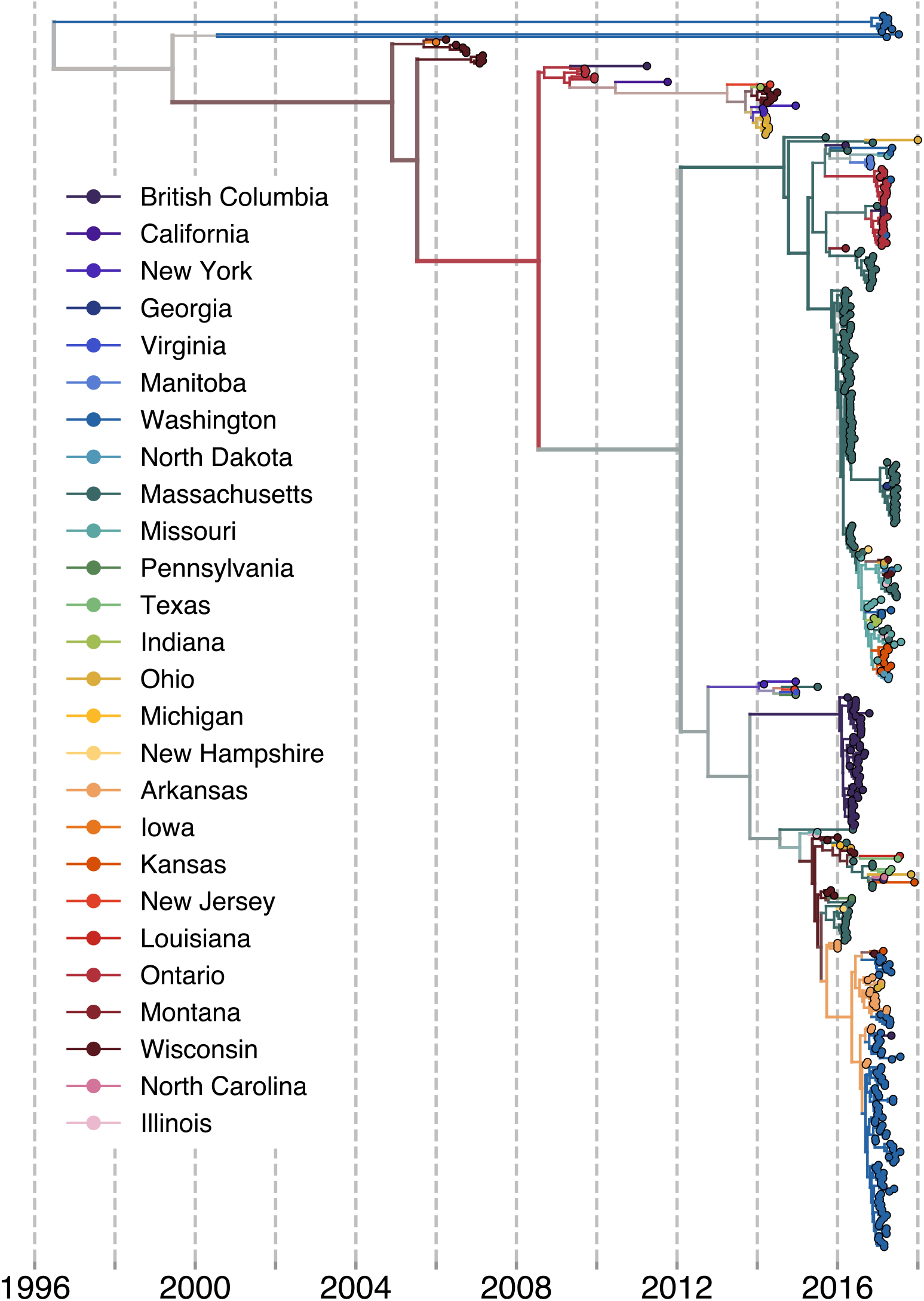
North American mumps outbreaks are related. We combined all publicly available North American mumps genomes and built a time-resolved phylogeny. Here, we display the maximum clade credibility tree, where each color represents a unique US state or Canadian province and the x-axis represents the collection date (for tips), or the inferred time to the most recent common ancestors (for internal nodes). We inferred geographic history using a discrete trait model. The color of each internal node represents the posterior probability of the inferred geographic location, where increasingly grey tone represents decreasing probability.

### Mumps was introduced into Washington multiple independent times

Estimating the number and timing of viral introductions is important for estimating epidemiologic parameters and evaluating surveillance networks, but is challenging with case count data alone. The Washington Department of Health had identified a single potential index case in October of 2016. To determine whether the genomic data supported a single introduction, we separated each introduction inferred in the maximum clade credibility tree and plotted each as its own transmission chain (**Fig. 3a**). We then enumerated the number of transitions into Washington in each tree in the posterior set, and plotted the distribution of Washington introductions consistent with the phylogeny (**Fig. 3b**).

**Figure 3:**
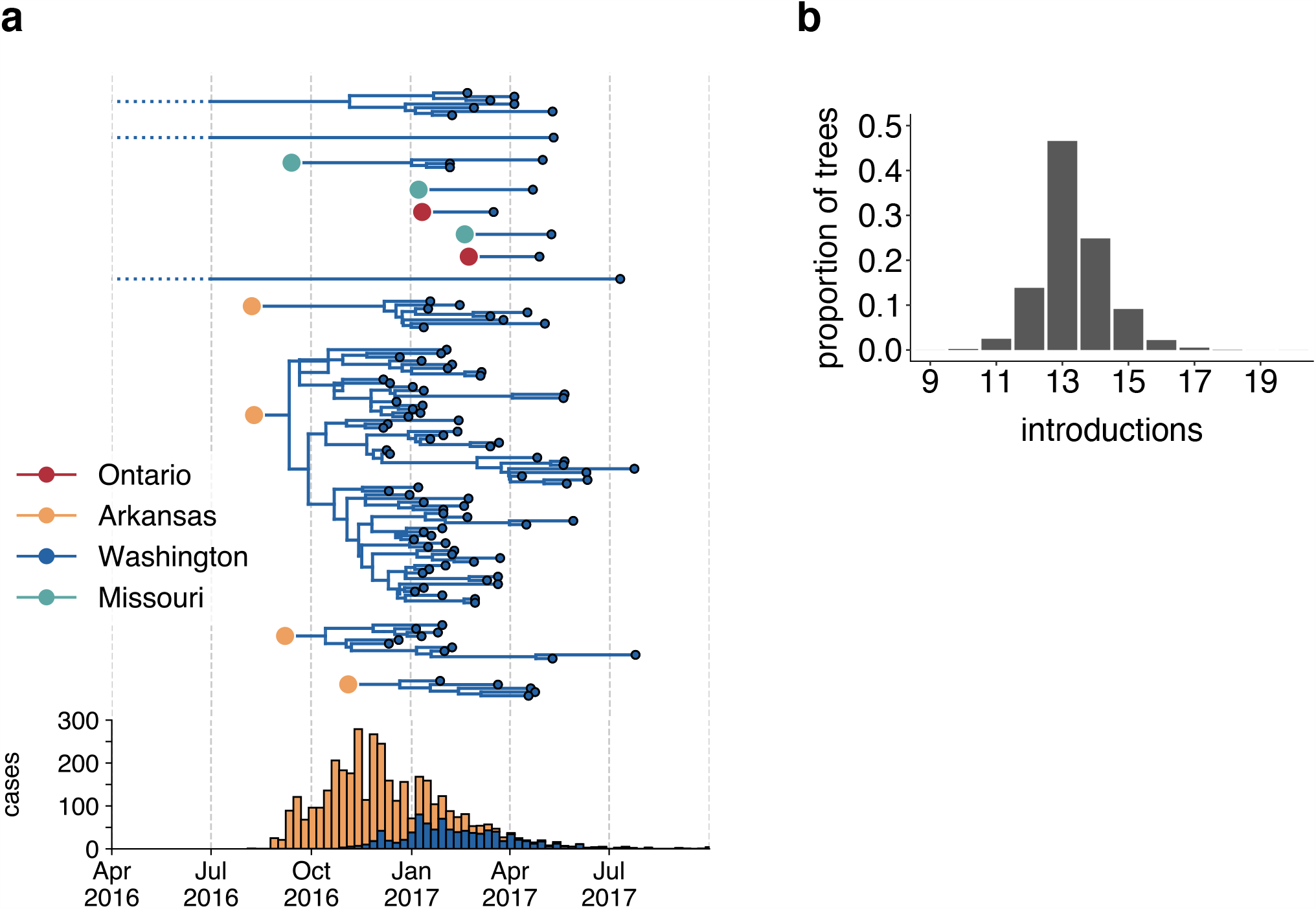
The mumps outbreak in Washington was seeded by approximately 13 introductions. We separated each introduction inferred on the maximum clade credibility tree (Figure 2) and plotted them independently. Large, colored dots represent the inferred geographic location that the Washington introduction was seeded from. Branches that extend earlier than July of 2016 are dotted to represent that transmission likely occurred via other, unsampled locations. For reference, the cumulative case counts from Arkansas and Washington are plotted below. **b**. For each tree in the posterior set, we inferred the number of introductions into Washington. We plot the proportion of trees in the posterior set in which that number of introductions was inferred.

Genomic data show that mumps was introduced into Washington State approximately 13 independent times (95% highest posterior density, HPD: 12 - 15), from geographically disparate locations (**Fig. 3**). Nine sampled tips descend from long branches (∼22 years), suggesting likely transmission from unsampled geographic locations. We infer introductions from Ontario and Missouri that each lead to 1-3 sampled cases (**Fig. 3b**), suggesting limited onward transmission following these introductions. In contrast, 4 introductions from Arkansas account for 92/110 sequenced cases, suggesting that these introductions led to more sustained chains of transmission following introduction (**Fig. 3b**). We refer to the largest cluster as the “primary outbreak clade,” and infer its introduction from Arkansas to Washington around August of 2016 (August 7, 2016, 95% HPD: July 11, 2016 to September 19, 2016, **Fig. 3b**), 3.5 months before Washington’s first reported case. These data reveal that what had appeared to be a single outbreak based on case surveillance data was in fact a series of multiple introductions, primarily from Arkansas, sparking overlapping and co-circulating transmission chains.

### SH gene sequencing is insufficient for fine-grained geographic inference

Mumps virus surveillance and genotyping relies on the SH gene (Centers for Disease Control and Prevention, 2019a), a short, 316 bp gene that is simple and rapid to sequence. To determine whether SH gene sequencing would have produced similar results, we built a divergence tree using our set of North American full genomes (**Supplemental Figure 2a**), then truncated that data to include only SH gene sequences (**Supplemental Figure 2b**). Almost all North American SH genes were identical, resulting in a single, large polytomy (**Supplemental Figure 2b**). This indicates that SH sequences lack sufficient resolution to elucidate fine-grained patterns of geographic spread, consistent with previous findings (Gouma et al., 2016; Wohl et al., 2020).

### Quantifying differences in transmission patterns within Washington

In both Arkansas and Washington, Marshallese individuals comprised over 50% of mumps cases, despite accounting for a much lower proportion of the population in both states. Phylogenetic reconstruction links the outbreaks in Washington and Arkansas, placing most sampled mumps genomes in Washington as descendant from Arkansas. We sought to investigate how mumps transmission may have differed within Marshallese and non-Marshallese communities within the same outbreak. Phylogenetic trees reflect the transmission process and can be used to quantify differences in transmission patterns among population groups. If transmission rates were distinct between Marshallese and non-Marshallese mumps cases, we expect the following: 1. Sequences from the high-transmitting group should be more frequently detected upstream in transmission chains. 2. Introductions seeded into the high-transmitting group should result in larger and more diverse clades in the tree. 3. The internal nodes of the phylogeny should be predominantly composed by members of the high-transmitting group, while members of the low-transmitting group should primarily be found at terminal nodes.

### Marshallese cases are enriched upstream in transmission chains

We developed a transmission metric to quantify whether Marshallese cases were enriched at the beginnings of successful transmission chains. We traverse the full genome divergence phylogeny (**Supplemental Figure 2a**) from root to tip. When we encounter a tip that lies on an internal node, we enumerate the number of tips that descend from its parent node. We then classify each tip in the phylogeny as either having descendants or not, and compare the proportion of tips with and without descendants among groups (**Fig. 4a**, see Methods for more details). Given our sampling proportion (110 sequences/889 total cases, ∼12%), we do not expect to have captured true parent/child infection pairs. Rather, we expect to have preferentially sampled long, successful transmission chains within the state. This allowed us to assess whether infections with particular attributes (community membership, vaccination status, age) are predictors for being upstream in these chains, and thus associated with sustained transmission. We evaluated the probability of having descendants in the tree as a function of vaccination status, age, and community status with logistic regression (see Methods for details and full model). Neither age nor vaccination status were significantly associated with the presence of downstream tips in the tree (**Table 1**). However, Marshallese cases were significantly more likely to have downstream descendants than non-Marshallese cases (odds ratio = 3.2, p = 0.00725, **Table 1**). While only 27% (14/52) of non-Marshallese tips were ancestral to downstream samples, 56% (32/57) of Marshallese tips had downstream descendents. These results suggest that community membership was a significant determinant of sustained transmission while controlling for vaccination status and age.

**Table 1:** Logistic regression results of probability that phylogeny nodes has descendants.

| Predictor variable | Estimated coefficient (standard error) | Odds ratio (95% CI) | p-value |
| --- | --- | --- | --- |
| Not up-to-date | -0.77 (0.69) | 0.46 (0.11, 1.70) | 0.26 |
| Vaccination status unknown | 0.79 (0.80) | 2.21 (0.59, 9.20) | 0.32 |
| Age | -1.2 (1.44) | 0.30 (0.016, 4.70) | 0.41 |
| Community status | 1.17 (0.43) | 3.21 (1.39, 7.69) | 0.0073 |

We evaluated the impact of vaccination status, age, and community membership on the probability that the phylogenetic node had descendants in the tree. Coefficients represent the increase in log odds of having descendants for the given predictor variable. Coefficients were exponentiated to produce odds ratios. We evaluated the impacts of having an unknown vaccination status, having a vaccination status that is not up-to-date, and being Marshallese as binary predictor variables. Age was evaluated as a continuous variable normalized such that values fall between 0 and 1.

**Figure 4:**
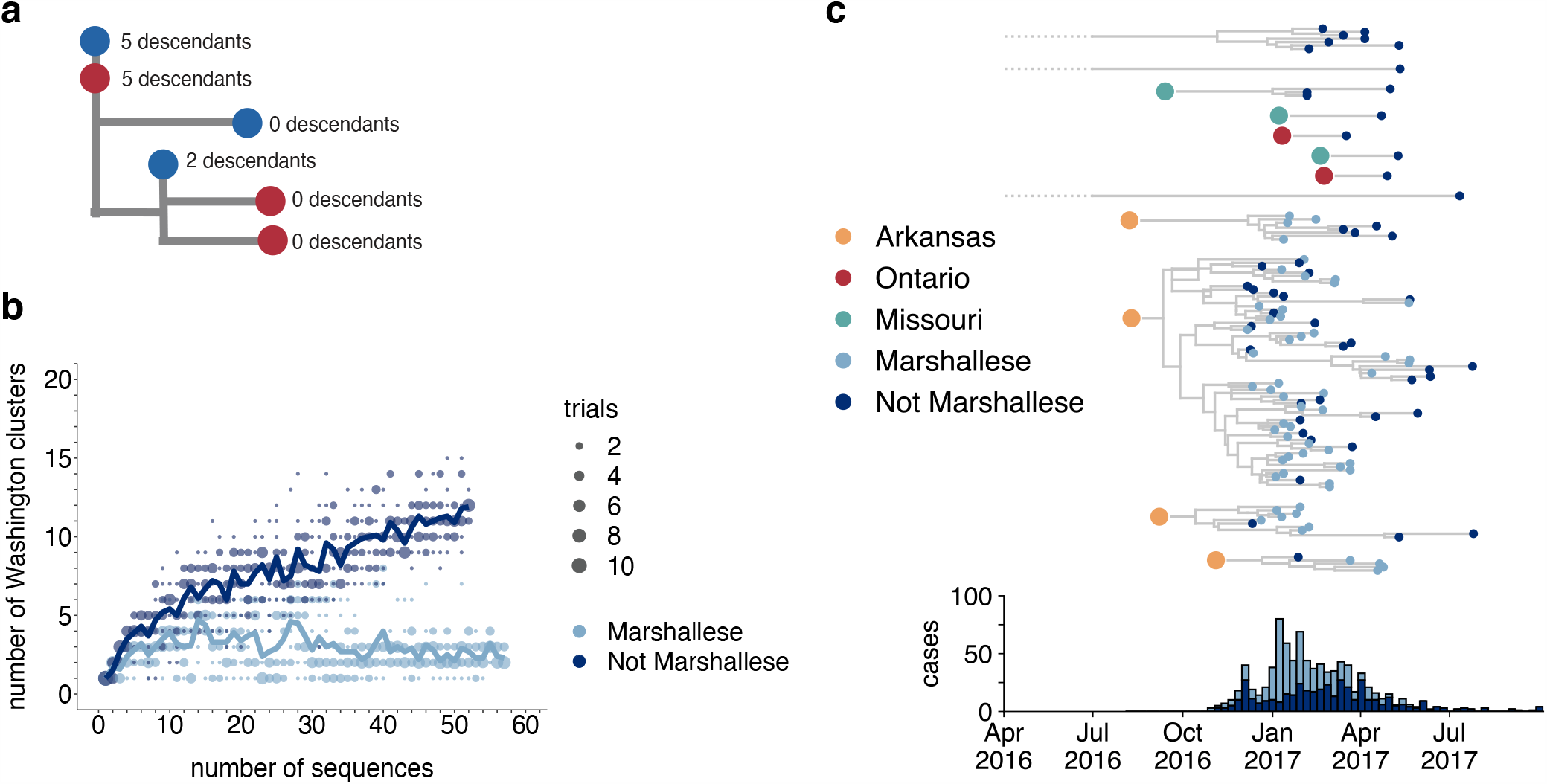
Transmission chains are longer within the Marshallese community. **a**. A schematic for quantifying tips that lie “upstream” in transmission chains. For tips that lie on an internal node, i.e., have a branch length of zero, we infer the number of child tips that descend from that tip’s parental node. For each tip in the example tree, the number of descendants we would infer is annotated alongside it. All tips that have a nonzero branch length are annotated as having 0 descendants. We can then compare whether sequences of particular groups (here, blue vs. red) are more likely to have descendants in the tree via logistic regression. **b**. We separated all Washington tips and classified them into Marshallese and not Marshallese. We then performed a rarefaction analysis and plot the number of inferred Washington clusters (y-axis) as a function of the number of sequences included in the analysis (x-axis). Dark blue represents not Marshallese sequences, and light blue represents Marshallese sequences. Each dot represents the number of trials in which that number of clusters was inferred, and the solid line represents the mean across trials. **c**. The exploded tree as shown in Figure 3a is shown, but tips are now colored by whether they represent Marshallese or non-Marshallese cases. For reference, the number of Washington cases (y-axis) is plotted over time (x-axis), where bar color represents whether those cases were Marshallese or not.

### Longer transmission chains are associated with community status

In the absence of recombination, closely-linked infections will cluster together on the tree, while unrelated infections should fall disparately on the tree, forming multiple smaller clusters. We inferred the number of Washington-associated clades in the tree as a function of whether sampled infections came from Marshallese or non-Marshallese individuals. Using the full North American phylogeny, we removed all Washington sequences and separated them into viruses sampled from cases noted as Marshallese or non-Marshallese. Then, separately for each group, we added sequences back into the tree one by one, until all sequences for that group had been added. For each number of sequences, we performed 10 independent trials (see Methods for complete details), and at each step, enumerated the number of inferred Washington clusters in the phylogeny. For comparison, we also grouped tips by vaccination status and repeated this analysis.

For tips from non-Marshallese individuals, the number of inferred clusters increases linearly as tips are added to the tree (**Fig. 4b**). This suggests that these infections are not directly related, and are not part of sustained transmission chains (**Fig. 4b**). In contrast, the number of inferred clusters for Marshallese tips stabilizes after ∼10 tips are added, even as almost 50 more sequences are added to the tree. This pattern likely arises because many Marshallese infections are part of the same long transmission chain, such that newly added tips nest within existing clusters. We do not observe similar differences among vaccination groups (**Supplemental Figure 3**). These findings are consistent with distinct patterns of transmission among Marshallese versus non-Marshallese cases: transmission among Marshallese individuals resulted in a small number of large clusters, while transmission among non-Marshallese individuals are generally the result of disparate introductions that generate shorter transmission chains.

We next separated each Washington introduction and colored each tip by community membership. Every introduction that was not seeded from Arkansas led to exclusively non-Marshallese infections, while introductions from Arkansas defined lineages that circulated for longer and were enriched with Marshallese tips (**Fig. 4c**). The primary outbreak clade is particularly enriched, containing 43 Marshallese tips and 26 non-Marshallese tips, hinting that transmission chains are longer when Marshallese cases are present in a cluster.

### Mumps transmitted efficiently within the Marshallese community

Internal nodes on a phylogeny represent ancestors to subsequently sampled tips, while terminal nodes represent viral infections that did not give rise to sampled progeny. If the mumps outbreak were primarily sustained by transmission within one group, the backbone of the phylogeny and the majority of internal nodes should be inferred as circulating in that group. We selected the 4 introductions that contained both Marshallese and non-Marshallese tips (**Fig. 4c**, the 4 Arkansas introductions), and reconstructed ancestral states along the phylogeny and migration/transmission rates between Marshallese and non-Marshallese groups using a structured coalescent model.

74/88 internal nodes were inferred to circulate within the Marshallese community with posterior probability of at least 0.95 (**Fig. 5a, b**). Movement of a lineage from the Marshallese deme into the non-Marshallese deme subsequently caused the lineage to die out quickly (**Fig. 5a**, dark blue branches). This suggests that transmission was overwhelmingly maintained within the Marshallese community, and that infections seeded into the non-Marshallese community did not sustain prolonged transmission chains. We estimate substantially more transmission from Marshallese to non-Marshallese groups than the opposite: within the primary outbreak clade, we estimate 29 transmission events from Marshallese to non-Marshallese groups (95% HPD: 21, 37), and only 6 (95% HPD: 0, 14) from non-Marshallese to Marshallese groups (**Fig. 5d**). This strongly suggests that transmission predominantly occurred in one direction: transmission events leading to non-Marshallese infections usually died out, and did not typically re-seed circulation within the Marshallese community. These results hold true regardless of migration rate prior (**Supplemental Figure 4**).

**Figure 5:**
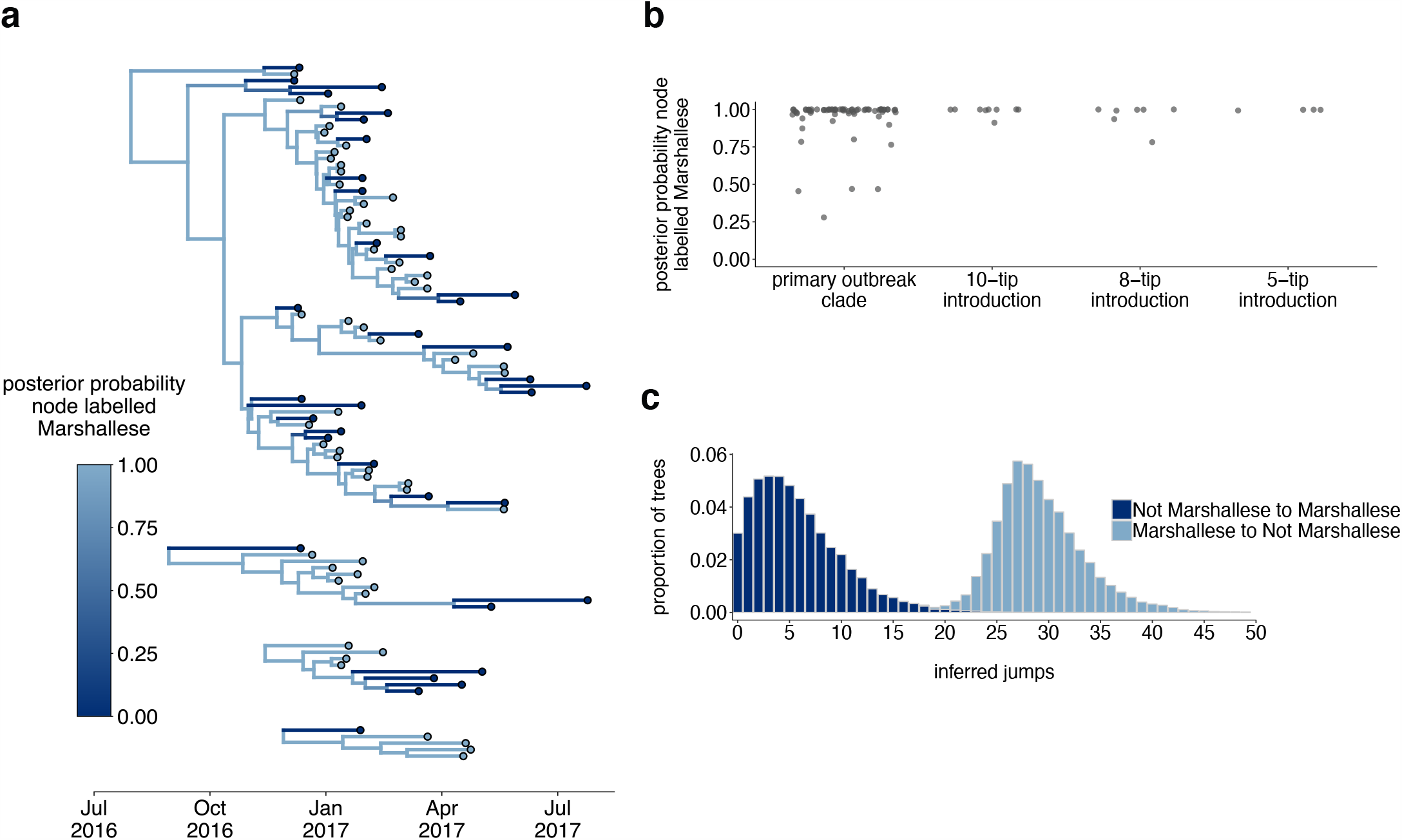
The Washington outbreak was sustained by transmission in the Marshallese community. **a**. Using the 4 Washington clusters that had a mixture of Marshallese and non-Marshallese cases, we inferred phylogenies using a structured coalescent model. Each group of sequences shared a clock model, migration model, and substitution model, but each topology was inferred separately, allowing us to incorporate information from all 4 clusters into the migration estimation. For each cluster, the maximum clade credibility tree is shown, where the color of each internal node represents the posterior probability that the node is Marshallese. **b**. For each internal node shown in panel a, we plot the posterior probability of that node being Marshallese. Across all 4 clusters, 74 out of 88 internal nodes (84%) are inferred as Marshallese with a posterior probability of at least 0.95. **c**. The posterior distribution of the number of “jumps” or transmission events from Marshallese to not Marshallese (light blue) and not Marshallese to Marshallese (dark blue) inferred for the primary outbreak clade.

To ensure that our results were not driven by unequal sampling within the analyzed clades, we generated 3 datasets in which the number of Marshallese and non-Marshallese tips were subsampled to be equal. For each of these 3 subsampled datasets, we ran 3 independent chains under the same model described above. Chains converged for 2 of the 3 subsampled datasets. In the converged chains, we recover very similar tree topologies (**Supplemental Figure 5a**) with equivalent phylogenetic reconstructions of lineage circulation within Marshallese and non-Marshallese demes. We also recovered maximum clade credibility trees in which the vast majority of the internal nodes are inferred to circulate within the Marshallese deme (**Supplemental Figure 5a**,**b**), confirming that our findings are robust to sampling, consistent with past observations of model performance (De Maio et al., 2015; Dudas et al., 2018; Vaughan et al., 2014).

The structured coalescent model requires both groups to be present in each cluster, excluding several small Washington introductions composed entirely of non-Marshallese tips (**Fig. 4c**). To account for this, we used all Washington genotype G sequences in our dataset and estimated a single tree using an approximate structured coalescent model (Müller et al., 2018). All Washington sequences were annotated as either Marshallese or not Marshallese. To provide a “source” population for the extensive diversity among our disparate Washington introductions, we also specified a third, unsampled deme, for which migration was only allowed to proceed outward. As above, we inferred very few non-Marshallese internal nodes (**Supplemental Figures 6 and 7**). All internal nodes in the primary outbreak group are inferred as Marshallese with high probability, while non-Marshallese cases are present as terminal nodes. We recovered support for a single non-Marshallese cluster, indicating limited sustained transmission in the non-Marshallese population.

Structured coalescent models infer the effective population size (*N*_*e*_) for each group, which reflects the number of infections necessary to generate the observed genetic diversity. Differences in *N*_*e*_ can result from different transmission rates or different numbers of infected individuals (Volz, 2012), and can therefore approximate differences in disease frequency between groups. While the total number of Marshallese and non-Marshallese cases reported through the public health surveillance system were similar (**Supplemental Table 2**), we estimate that *N*_*e*_ for the non-Marshallese group is approximately 3 times higher than that of the Marshallese group. Assuming the same number of infected individuals in each group, lower *N*_*e*_’s suggest higher transmission rates (Volz, 2012), suggesting more transmission within the Marshallese deme. Taken together, our results suggest that the outbreak was primarily sustained by transmission within the Marshallese community. While this transmission sometimes spilled over into the non-Marshallese community, transmission was generally not as successful there and died out, resulting in short, terminal transmission chains.

### Viruses infecting vaccination groups individuals are genetically similar

Although only 9.7% of reported mumps cases in Washington were not up-to-date for mumps vaccination, infection of these individuals could have disproportionately impacted transmission in the state. Emergence of an antigenically novel strain of mumps could also allow infection of previously vaccinated individuals, and result in different virus lineages infecting individuals in different vaccination categories. We colored the tips of all Washington cases in our phylogeny to represent whether they were derived from individuals who were up-to-date, not up-to-date, or whose vaccination status was unknown. Mirroring overall vaccination coverage in Washington, the vast majority of samples in our dataset were from up-to-date individuals. The not up-to-date individuals present in our dataset are dispersed throughout the phylogeny and do not cluster together (**Fig. 6**), suggesting that there is no genetic difference between viruses infecting individuals with different vaccination statuses.

**Figure 6:**
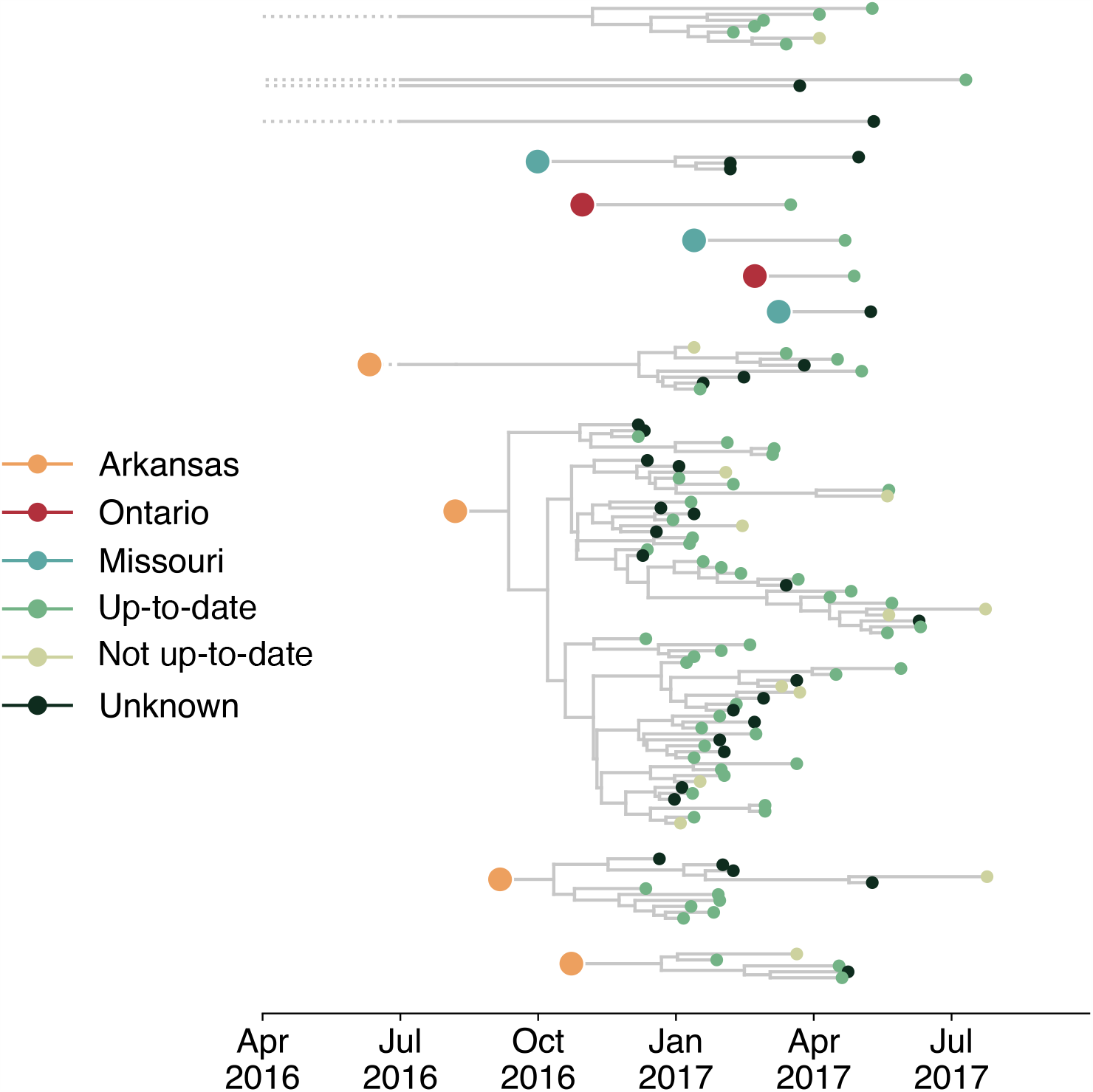
Individuals in different vaccination groups are infected by genetically similar viruses. The exploded tree as shown in Figure 3a is shown, but tips are now colored by whether they represent cases from individuals who are up-to-date for mumps vaccination, not up-to-date, or cases for which vaccination status was unknown. The color of the large dot represents the inferred geographic location from which the Washington introduction was seeded.

## Discussion

The resurgence of mumps in North America has ushered renewed attention towards understanding post-vaccine era mumps transmission. While many studies have used phylodynamic approaches to elucidate viral patterns of geographic spread (Dudas et al., 2017; Gouma et al., 2016; Grubaugh et al., 2017; Stapleton et al., 2019), using genomics to distinguish transmission patterns among epidemiologically distinct groups is novel. We employ a phylogenetic method (Vaughan et al., 2014) traditionally applied to geography that is robust to sampling bias (Dudas et al., 2018) to investigate drivers of mumps transmission in Washington. We show that the Washington State outbreak was fueled by approximately 13 independent introductions, primarily from Arkansas, leading to multiple co-circulating transmission chains.

Within Washington, transmission was more efficient within the Marshallese community. Marshallese individuals were more often sampled at the beginnings of transmission chains, contributed to longer transmission chains on average, and were overwhelmingly enriched on internal nodes within the phylogeny. We found no support for age or vaccination status as critical determinants of transmission in our outbreak, consistent with epidemiologic findings. Our data suggest that social networks can be critical determinants of mumps transmission. Future work exploring how social and economic disparities may amplify respiratory disease transmission will be necessary for updating outbreak mitigation and prevention strategies. By combining detailed metadata, novel metrics of transmission in the tree, and robust controls for sampling, we provide a framework for investigating source-sink dynamics that are readily applicable to other viral pathogens.

Our results highlight the utility of genomic data to clarify epidemiologic hypotheses. While genomic data and epidemiologic investigation (including case interviews and contact follow up) suggested an Arkansas introduction as the Washington outbreak’s primary origin, sequence data revealed repeated and ongoing introductions into Washington, similar to patterns observed in Massachusetts, and the Netherlands (Gouma et al., 2016; Wohl et al., 2020). We also find widespread geographic mixing across the phylogeny, consistent with investigations from the US (Wohl et al., 2020), Canada (Stapleton et al., 2019), and Europe (Gavilán et al., 2018; Gouma et al., 2016). Like others (Gouma et al., 2016; Wohl et al., 2020), we confirm that SH genotyping alone is insufficient for fine-grained resolution of geographic transmission patterns. While CDC guidelines currently recommend SH-based genotyping specifically for tracking transmission pathways (Clemmons et al., 2020), building public health capacity for full-genome sequencing may provide better resolution for resolving local mumps transmission patterns.

Despite similar numbers of Marshallese and non-Marshallese cases, we find that mumps did not transmit efficiently among the general Washington populace. Washington State uses a passive surveillance system for mumps detection and case acquisition, which are known to frequently result in underreporting. Because the WA Department of Health did not perform active mump surveillance, it is difficult to assess whether different epidemiologic groups have different likelihoods of being sampled. Marshallese individuals are less likely to seek healthcare (Towne et al., 2020), which may have resulted in particularly high rates of underreporting in this group. Although statewide vaccine coverage is high, the risk for acquiring and transmitting mumps is clearly not equal across all Washington population groups. Provision of an outbreak dose of mumps-containing vaccine to high-risk groups may therefore be especially effective for limiting mumps transmission in future outbreaks. Others have reported success in using outbreak dose mumps vaccinations to reduce mumps transmission on college campuses (Cardemil et al., 2017; Shah et al., 2018) and in the US army (Arday et al., 1989; Eick et al., 2008; Green, 2006; Kelley et al., 1991), and the CDC currently recommends providing outbreak vaccine doses to individuals with increased risk due to an outbreak (Marlow et al., 2020). Future work to quantify the interplay between contact rates and vaccine-induced immunity among different age and risk groups should be used to guide updated vaccine recommendations.

Recent research has focused on identifying groups at risk for mumps infection due to their age (Lewnard and Grad, 2018), with less attention to other factors that may make populations vulnerable. In the 2016/017 Washington State outbreak, Marshallese status was a crucial predictor of transmission risk. Mumps has historically caused outbreaks in communities with strong, interconnected contact patterns (Barskey et al., 2012; Fields et al., 2019; Nelson et al., 2013), and in dense housing environments (Snijders et al., 2012). While a combination of waning immunity and dense housing settings make college campuses ideal for mumps outbreaks, the Washington and Arkansas outbreaks show that populations other than young adults are at risk. Soliciting feedback from a Marshallese community health advocate allowed us to contextualize our genomic results with the lived experience of individuals most heavily affected during the outbreak and to identify reasonable hypotheses to explain the efficient transmission we observe. Based on these interviews and previously published studies, we speculate that within the Marshallese community, a combination of factors likely led to a high force of infection.

Multigenerational living is common in the Marshallese community (Fields et al., 2019), and Marshallese households tend to be larger on average (average household size = 5.28 (Harris and Jones, 2005), average household size for entire US populace = 2.52 (US Census Bureau, n.d.)). Having more household contacts may have facilitated a greater number and higher intensity of interactions among individuals, allowing the force of infection to overcome pre-existing immunity. It is also possible that infection intensity within the Marshallese community was exacerbated by low rates of insurance coverage and poor access to healthcare (McElfish et al., 2017; Towne et al., 2020), hesitancy to seek medical care (Williams and Hampton, 2005), and health disparities stemming from US occupation, nuclear testing, and exclusion from healthcare services. As part of reparations for US nuclear testing, the US signed the Compact of Free Association Treaty (COFA)(Congress, 2003) with the Marshall Islands in 1989, permitting Marshallese residents to live and work in the US without visas. However, eligibility for Medicaid was revoked for COFA immigrants in 1996, and US-residing Marshallese remain economically disadvantaged and under-insured (McElfish, 2016; McElfish et al., 2017, 2015). The passage of the Affordable Care Act (ACA) has not ameliorated these issues. Interviews with US-residing Marshallese note confusion among ACA staff regarding the legal status of COFA recipients, leading to drawn out enrollment processes that often leave individuals uninsured, frustrated (McElfish et al., 2016), and far less likely to access care (Towne et al., 2020). A study of healthcare-seeking behavior among patients with diabetes showed that while multiple factors contribute to forgone care in the US populace, 77% of surveyed Marshallese individuals reported recent forgeone care and cited lack of insurance as the primary reason (Towne et al., 2020). Marshallese trust in US medical institutions was seriously undermined by the unconsented use of Marshallese individuals for experiments on health impacts of nuclear exposure, with effects lingering today (Barker, 2012). Banked historical samples confirm uptake of radioactive materials in Marshallese inhabitants of affected Islands (Simon et al., 2010), but there has been limited published data on long-term health impacts of nuclear exposure, and significant concern remains within the community (Bordner et al., 2016). Finally, when Marshallese individuals do access care, they report experiencing disdain from healthcare workers (Duke, 2017) and sub-optimal care (McElfish et al., 2016). Interviews with medical workers show that blame for poor Marshallese health outcomes is sometimes placed on host genetics or cultural practices (Duke, 2017), poor health literacy (McElfish et al., 2018), or choosing to delay care (McElfish et al., 2018), with less consideration given to how the economic and legal impacts of US occupation affect the health of Marshallese individuals. These factors compound, and Marshallese individuals report hesitation to seek medical care, even when sick (McElfish et al., 2016). Hesitancy to seek care could have contributed to mumps transmission if sick individuals were primarily cared for at home without knowledge of or the ability to implement community-isolation protocols.

Our findings highlight that social networks can be the primary risk factor for a respiratory virus outbreak, even when a vaccine is effective and widely used. This finding is especially pertinent as SARS-CoV2 continues to disproportionately impact populations who live and work in high-risk settings, including the Marshallese (Center et al., 2020; McElfish et al., 2021), and for whom vaccine licensure and distribution alone may not be a panacea. The passing of federal legislation remedying the exclusion of Marshallese individuals from Medicaid access (Hirono, 2019) in December 2020 marks an important step toward improving healthcare access for US-residing Marshallese. Future work to evaluate whether this change improves Marshallese access to healthcare and mitigates increased disease risk will be crucial follow-up. Future work should also explore whether nuclear exposure has impacted Marshallese immune function and susceptibility to infectious disease. Our findings demonstrate the importance of expanding our understanding of populations at risk for mumps re-emergence, so that rapid and comprehensive outbreak response strategies can be implemented to mitigate negative health impacts for all affected communities. Finally, future work to disentangle the complex interplay between healthcare access, social and economic disparity, and respiratory virus risk will be essential for mitigating health impacts of mumps and other respiratory viruses.

## Materials and Methods

### Data and Code Availability

All code used to analyze data, input files for BEAST, and all code used to generate figures for this manuscript are publicly available at https://github.com/blab/mumps-wa-phylodynamics. Raw FASTQ files with human reads removed are available under SRA project number PRJNA641715. All protocols for generating sequence data as well as the consensus genomes are available at https://github.com/blab/mumps-seq. Consensus genomes have also been deposited to Genbank under accessions MT859507-MT859672.

### Community feedback

In order to ensure that this study was faithful to the experience of the Marshallese community in Washington State, we sought paid consultation from a local Marshallese community health advocate. We conducted video and telephone interviews to directly address the impacts of mumps transmission on the Marshallese community, community healthcare goals and priorities, and the impacts of the mumps outbreak on stigmatization. This feedback informed what is being presented herein, provided crucial context for understanding mumps transmission, and allowed us to work with the community to determine how best to discuss Marshallese involvement in the outbreak.

### Mumps surveillance in Washington State

Mumps is a notifiable condition in Washington State. Therefore, per the Washington Administrative Code (WAC), as specified in WAC Chapter 246-101(Washington State Legislature, 2014), healthcare providers, healthcare facilities, and laboratories must report cases of mumps or possible mumps to the local health jurisdiction (LHJ) of the patient’s residence. LHJ staff initiate case investigations and facilitate optimal collection and testing of diagnostic specimens. Buccal swabs and urine are acceptable specimens for real-time reverse transcription polymerase chain reaction (qRT-PCR), a preferred diagnostic test for mumps. Most mumps rRT-PCR tests for Washington State residents are performed at the Washington State Public Health Laboratories, where all positive specimens are archived.

Individuals testing positive for mumps ribonucleic acid (RNA) by qRT-PCR are classified as confirmed mumps cases if they have a clinically-compatible illness (i.e., an illness involving parotitis or other salivary gland swelling lasting at least 2 days, aseptic meningitis, encephalitis, hearing loss, orchitis, oophoritis, mastitis, or pancreatitis). During case investigations, case-patients or their proxies are interviewed. Information about demographics, illness characteristics, vaccination history, and potential for exposure to and transmission of mumps are solicited from each case-patient. In concordance with CDC guidelines (Centers for Disease Control and Prevention, 2019b), only vaccine doses for which there was written documentation with the date of vaccine receipt were considered valid. Individuals for which such documentation could not be provided were classified as having an unknown vaccination status. For individuals with documented vaccine doses, they were further characterized as up-to-date or not up-to-date based on their age. The Washington State Department of Health (DOH) receives, organizes, performs quality control on, and analyzes data from, LHJ case reports and supports investigations upon request.

### Sample collection and IRB approval

This study was approved by the Fred Hutchinson Cancer Research Center (FHCRC) Institutional Review Board (IR File #: 6007-944) and by the Washington State Institutional Review Board, and classified as not involving human subjects. Samples were selected for sequencing to maximize temporal and epidemiologic breadth and to ensure successful sequencing. As such, samples were chosen based on the date of sample collection, the PCR cycle threshold (Ct), case vaccination status, and community status (Marshallese or non-Marshallese). Samples were selected for sequencing in 2 batches. In the first, samples were selected based on covering a wide geographic range within Washington, a full range of dates covering the outbreak, and having a Ct value < 36. This initial sampling regime resulted in a sample set skewed slightly towards samples from Marshallese individuals. To ensure that the proportion of samples in our data closely matched the distribution of cases in the outbreak, we then selected a second batch of samples using the same criteria as above, but excluded samples from Marshallese individuals. We then randomly sampled an additional 30 samples from non-Marshallese individuals. This sampling regime resulted in a dataset that closely mirrors the distribution of metadata categories in the outbreak overall. All metadata, including case vaccination status, were transferred from WA DOH to FHCRC in a de-identified form.

We also sequenced an additional set of 56 samples collected in Wisconsin, Ohio, Missouri, Alabama, and North Carolina provided by the Wisconsin State Laboratory of Hygiene. 10 of these samples were collected in Wisconsin during the 2006/2007 Midwestern college campus outbreaks, 6 samples were collected in 2014, and the rest were collected between 2016 and 2018. For these samples, we received metadata describing sample Ct value and date of collection. All metadata were received by FHCRC in de-identified form.

### Viral RNA extraction, cDNA synthesis, and amplicon generation

Viral RNA was extracted from buccal swabs using either the QiAmp Viral RNA Mini Kit (Qiagen, Valencia, CA, USA) or the Roche MagNA Pure 96 DNA and viral NA small volume kit (Roche, Basel, Switzerland). For samples extracted with the QiAmp Viral RNA Mini Kit, 500 μl of buccal swab fluid was spun at 5000 x g for 5 minutes at 4°C to pellet host cells. The supernatant was then removed and centrifuged at 14,000 rpm for 90 minutes at 4°C to pellet virions. Excess fluid was discarded, and the peletted virions were resuspended in 150-200 μl of fluid. Resuspended viral particles were then used as input to the QiAmp Viral RNA Mini Kit (Qiagen, Valencia, CA, USA), following manufacturer’s instructions, and eluting in 30 μl of buffer AVE. For extraction with the MagNA Pure, we followed manufacturer’s instructions.

cDNA was generated with the Protoscript II First strand synthesis kit (New England Biolabs, Ipswich MD, USA), using 8 μl of vRNA as input and priming with 2 μl of random hexamers. vRNA and primers were incubated at 65°C for 5 minutes. Following this incubation, 10 μl of Protoscript II reaction mix (2x) and 2 μl of Protoscript II enzyme mix (10x) were added to each reaction and incubated at 25°C for 5 minutes, then 42°C for 1 hour, followed by a final inactivation step at 80°C for 5 minutes. To amplify the full mumps genomes, we used Primal Scheme (http://primal.zibraproject.org/) to design overlapping, ∼1500 base pair amplicons spanning the entirety of the mumps virus genome, where each tiled set of primes overlapped by ∼100 base pairs. Primers are listed below.

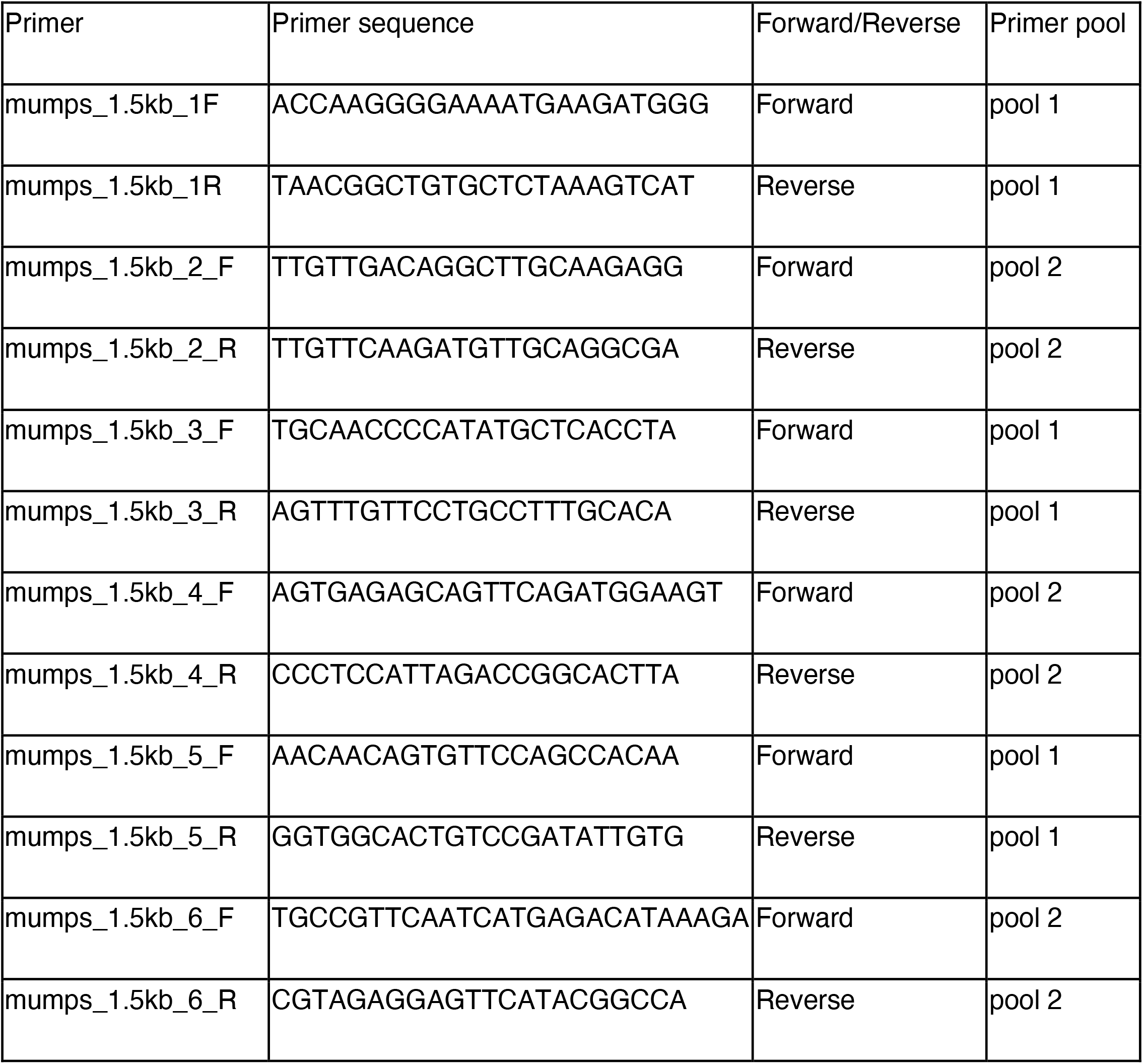
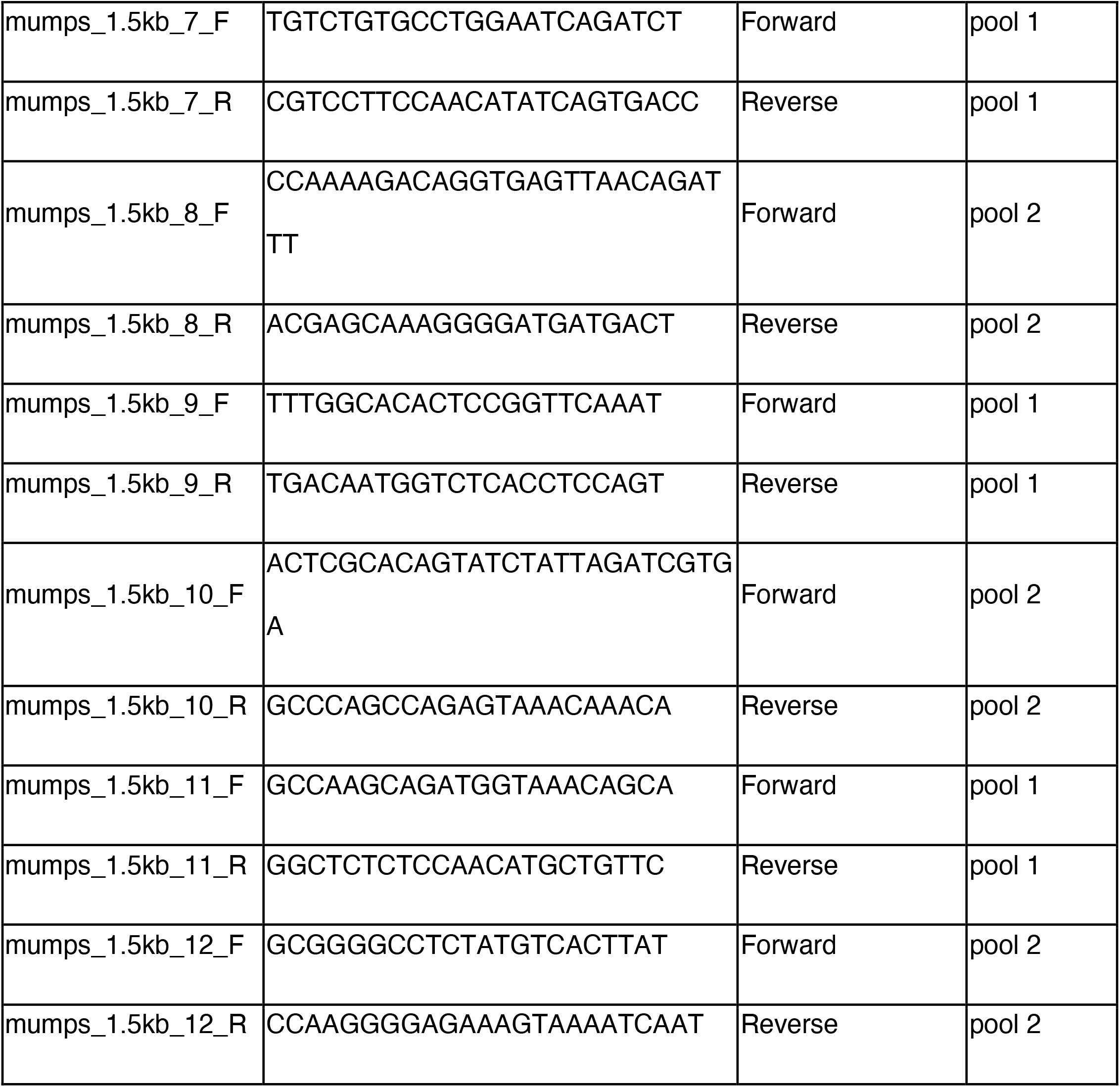

Primers were pooled into 2 pools as follows: the first contained primer pairs 1, 3, 5, 7, 9, and 11, all pooled at 10 uM. The second pool contained primer pairs 2, 4, 6, 8, 10, and 12. All primers in pool 2 were pooled at 10 uM, except for primer pair 4, which was added at a 20 uM concentration.

PCR was performed with the Q5 Hotstart DNA polymerase (New England Biolabs, Ipswich, MD, USA), using 11.75 μl of nuclease-free water, 5 μl of Q5 reaction buffer, 0.5 μl of 10 mM dNTPs, 0.25 μl, 2.5 of pooled primers, and 5 μl of cDNA. Amplicons were generated with the following PCR cycling conditions: 98°C for 30 seconds, followed by 30 cycles of: 98°C for 15 seconds, then 67°C for 5 minutes. Cycling was concluded with a 10°C hold. PCR products were run on a 1% agarose gel, and bands were cut out and purified using the QiAquick gel extraction kit (Qiagen, Valencia, CA, USA), following the manufacturer’s protocol. All optional steps were performed, and the final product was eluted in 30 *µ*l of buffer EB. For samples extracted on the MagNA Pure, amplicons were cleaned using a 1x bead cleanup with Ampure XP beads. Final cleaned amplicons were quantified using the Qubit dsDNA HS Assay kit (Thermo Fisher, Waltham, MA, USA).

### Library preparation and sequencing

For each sample, pool 1 and pool 2 amplicons were combined in equimolar concentrations to a total of 0.5 ng in 2.5 *µ*l. Libraries were prepared using the Nextera XT DNA Library Prep Kit (Illumina, San Diego, CA, USA), following manufacturer’s instructions, but with reagent volumes halved for each step, for the majority of samples in our dataset. For samples processed in our last sequencing run, several samples had higher Ct values. We therefore chose to process these samples using the standard 1x reagent volumes for the library preparation step. All libraries were purified using Ampure XP beads (Beckman Coulter, Brea, CA, USA), using a 0.6x cleanup, a 1x cleanup, and a final 0.7x cleanup. At each step, beads were washed twice with 200-400 *µ*l of 70% ethanol. The final product was eluted off the beads with 10 *µ*l of buffer EB. Tagmentation products were quantified with the Qubit dsDNA HS Assay kit (Thermo Fisher, Waltham, MA, USA), and run on a Tapestation with the TapeStation HighSense D5K assay (Agilent, Santa Clara, CA, USA) to determine the average fragment length. All but 8 samples and negatives were pooled together in 6 nM libraries and run on 300 bp x 300 bp v3 kits on the Illumina MiSeq, with a 1% spike-in of PhiX. The remaining 8 samples (MuVs/Washington.USA/1.17/FH77[G], MuVs/Washington.USA/12.17/FH78[G], MuVs/Washington.USA/16.17/FH79[G], MuVs/Washington.USA/19.17/FH80[G], MuVs/Washington.USA/20.17/FH81[G], MuVs/Washington.USA/20.17/FH82[G], MuVs/Washington.USA/29.17/FH83[G], and MuVs/Washington.USA/2.17/FH84[G]) were pooled to a 1.2 nM library, and run as a 50 pM library with 2% PhiX on the Illumina iSeq, with a 151 bp x 151 bp v3 kit.

### Negative controls

A negative control (nuclease-free water) was run for each viral RNA extraction, reverse transcription reaction, and for each pool for each PCR reaction. These negative controls were carried through the library preparation process and sequenced alongside actual samples. Any samples whose negative controls from any step in the process resulted in >10x mumps genome coverage were re-extracted and sequenced.

### Bioinformatic processing of sequencing reads

Human reads were removed from raw FASTQ files by mapping to the human reference genome GRCH38 with bowtie2 (Langmead and Salzberg, 2012) version 2.3.2 (http://bowtie-bio.sourceforge.net/bowtie2/index.shtml). Reads that did not map to the human genome were output to separate FASTQ files and used for all subsequent analyses. Illumina data was analyzed using the pipeline described in detail at https://github.com/lmoncla/illumina_pipeline. Briefly, raw FASTQ files were trimmed using Trimmomatic (Bolger et al., 2014) (http://www.usadellab.org/cms/?page=trimmomatic), trimming in sliding windows of 5 base pairs and requiring a minimum Q-score of 30. Reads that were trimmed to a length of <100 base pairs were discarded. Trimming was performed with the following command: java -jar Trimmomatic-0.36/trimmomatic-0.36.jar SE input.fastq output.fastq SLIDINGWINDOW:5:30 MINLEN:100. Trimmed reads were mapped to a consensus sequence from Massachusetts (Genbank accession: MF965301) using bowtie2(Langmead and Salzberg, 2012) version 2.3.2 (http://bowtie-bio.sourceforge.net/bowtie2/index.shtml), using the following command: bowtie2 - x reference_sequence.fasta -U read1.trimmed.fastq,read2.trimmed.fastq -S output.sam --local. We selected this Massachusetts sequence as an initial reference sequence because at the time, it represented one of the only available genomes of a genotype G mumps virus that had been sampled during a US outbreak in 2016. Mapped reads were imported into Geneious (https://www.geneious.com/) for visual inspection and consensus calling. To avoid issues with mapping to an improper reference sequence, we then remapped each sample’s trimmed FASTQ files to its own consensus sequence. These bam files were again manually inspected in Geneious, and a final consensus sequence was called, with nucleotide sites with <20x coverage output as an ambiguous nucleotide (“N”). All genomes with >50% Ns were discarded. In total, we generated 140 genomes with at least 80% non-N bases, and 26 genomes with 50-80% non-N bases. Our median completeness (percent of bases that are not Ns) across the dataset is 90%. All genomes used in these analyses are available at https://github.com/blab/mumps-seq/tree/master/data.

### Dataset curation and maximum likelihood divergence tree generation

We downloaded all currently available (as of June 2020), complete mumps genomes from North America from the NIAID Virus Pathogen Database and Analysis Resource (ViPR) (Pickett et al., 2012) through http://www.viprbrc.org/. We also obtained mumps genomes from British Columbia, Ontario, and Arkansas. We obtained written permission from sequence authors for any sequence that had not previously been published on. In total, this dataset includes 437 full mumps genomes from North America. Sequences and metadata were cleaned and organized using fauna, a database system that is part of the Nextstrain platform. Sequences were processed using Nextstrain’s augur software (Hadfield et al., 2018), and filtered to include only those with at least 8,000 bases and were sampled in North America in 2006 or later. Genomes were aligned with MAFFT (Katoh et al., 2002), and trimmed to the reference sequence (MuV/Gabon/13/2[G], GenBank accession: KM597072). We inferred a maximum likelihood phylogeny using IQTREE (Nguyen et al., 2015) with a GTR nucleotide substitution model, and inferred a molecular clock and temporally-resolved phylogeny using TreeTime (Sagulenko et al., 2018). Sequences with an estimated clock rate that deviated from the other sequences by >4 times the interquartile distance were removed from subsequent analysis. We inferred the root-to-tip distance with TempEst version 1.5.1 (Rambaut et al., 2016) with the best fitting root by the heuristic residual mean squared function.

### Phylogenetic analysis of full North American mumps genomes

Using the same set of genome sequences used for divergence tree estimation, we aligned sequences with MAFFT and inferred time-resolved phylogenies in BEAST version 1.8.4 (Drummond et al., 2012). We used a skygrid population size prior with 100 bins, and a skygrid cutoff of 25 years, allowing us to estimate 4 population sizes each year. We used an HKY nucleotide substitution model with 4 gamma rate categories, and a strict clock with a CTMC prior. We used a discrete trait model (Lemey et al., 2009) and estimated migration rates using BSSVS and ancestral states with 27 geographic locations. Here, “state” refers to the inferred ancestral identity of an internal node, where the inferred identity could be any of the 27 geographic locations (US states and Canadian provinces) in the dataset. For the prior on non-zero rates for BSSVS, we specified an exponential distribution with a mean of 26. As a prior on each pairwise migration rate, we used an exponential distribution with mean 1. All other priors were left at default values. We ran this analysis for 100 million steps, sampling every 10,000, and removed the first 10% of sampled states as burnin. A maximum clade credibility tree was summarized with TreeAnnotator, using the mean heights option. All tree plotting was performed with baltic (https://github.com/evogytis/baltic). Input XML files and output results are available at https://github.com/blab/mumps-wa-phylodynamics/tree/master/phylogeography.

### Testing for descendants in divergence trees

To determine whether specific groups were more likely to be found at the beginnings of transmission chains than other groups, we developed a statistic to quantify transmission in the tree. Using the tree JSON output from the Nextstrain pipeline (Hadfield et al., 2018), we traversed the tree from root to tip. For each tip that lay on an internal node, i.e., had a branch length of nearly 0 (< 1 x 10^−16^), we counted the number of descendants. We collapsed very small branches (those with branch lengths less 1 x 10^−16^) to obtain polytomies. We then classified tips as either having descendants (i.e., the number of descendents was > 0) or not having descendants. Here, we define a “descendant” as a tip that occurs in any downstream portion of the tree, i.e., it falls along the same lineage but to the right of the parent tip. A diagram of what we classify as “descendant tips” is shown in Figure 4a. The probability of having descendants was evaluated as a function of community status, age, and vaccination status with logistic regression as described below.

For each Washington tip in the tree, we classified it as either having descendants (coded as a 1) or not having descendants (coded as 0). For each tip, we coded it’s corresponding age, vaccination status, and community membership as a predictor variable input into a logistic regression model. We coded these attributes as follows: For community membership, non-Marshallese tips were coded as 0 and Marshallese tips were coded as 1. Age was coded as a single, continuous variable. In our dataset, there were 3 classifications for vaccination status: up-to-date, not up-to-date, and unknown vaccination status. According to the Advisory Committee on Immunization Practices (ACIP) (McLean et al., 2013), individuals aged 5-18 had to have received both recommended doses of mumps-containing vaccine, children aged 15 months to 5 years required 1 dose of mumps-containing vaccine, and adults over 18 had to have received at least 1 dose of mumps-containing vaccine to be classified as up-to-date for mumps vaccination. Individuals under 15 months are considered up-to-date without any doses of mumps-containing vaccine. Not up-to-date individuals are those with a known vaccination status who did not qualify under criteria to be classified as up-to-date. Individuals who could not provide documentation regarding their MMR vaccination history were considered to have “unknown” vaccination status. Individuals with “known” vaccination status could either be fully up-to-date, undervaccinated, or unvaccinated. To ensure that we measured the effect of vaccination among individuals who knew their vaccination status, we coded vaccination information using two dummy variables in our logistic regression, one signifying whether vaccination status was known or not, and one indicating whether vaccination was up-to-date or not. We then fit a logistic regression model to this data using the glm package in R (https://www.rdocumentation.org/packages/stats/versions/3.6.2/topics/glm), specifying a binomial model as

Pr(having descendants) ∼ *β*_0_ + *β*_1_*x*_1_ + *β*_2_*x*_2_ + *β*_3_*x*_3_ + *β*_4_*x*_*4*_, where *x*_1_ represents 0 or 1 value for member of Marshallese community (Not Marshallese coded as a 0, Marshallese coded as a 1), *x*_2_ represents the numeric value of age, *x*_3_ represents 0 or 1 value for whether vaccination status is unknown (having a known vaccination status coded as a 0, having an unknown vaccination status coded as a 1) and *x*_4_ represents 0 or 1 value for whether vaccination status is up-to-date (up-to-date coded as a 0 and not up-to-date coded as a 1). Under this formulation, an individual with unknown vaccination status would be coded as *x*_3_=1, *x*_4_=0, an individual who is up-to-date would be coded as *x*_3_=0, *x*_4_=0, and an individual who is not up-to-date is coded as *x*_3_=0, *x*_4_=1. This encoding allows us to evaluate the effects of having an unknown vaccination status and a vaccination status that is not up-to-date. Age was normalized such that values fall between 0 and 1 by: (*x*_2_ - minimum age in dataset) / (maximum age in dataset - minimum age in dataset).

P-values were assigned via a Wald test, and inferred coefficients were exponentiated to return odds ratios. All code used to parse the divergence tree and formulate and fit the regression model are available at https://github.com/blab/mumps-wa-phylodynamics/blob/master/divergence-tree-analyses/Regression-analysis-on-descendants-in-divergence-tree.ipynb.

### Rarefaction analysis to estimate transmission clusters

Using the full set of North American mumps sequences, we designated all non-Washington North American sequences as “background” sequences. We then separated Washington sequences into Marshallese tips (57 total sequences) and non-Marshallese tips (52 total sequences). For this analysis, we excluded the genotype K sequence in our dataset due to its extreme divergence from other viruses sampled in Washington, which were all genotype G. For each group (Marshallese vs. non-Marshallese), we then generated subsampled datasets comprised of a random sample of 1 to *n* sequences, where *n* is the number of total sequences available for that group. For each number of sequences, we performed 10 independent subsampling trials. Subsampling was performed without replacement. So, for community members, we generated 10 datasets in which 1 community member sequence was sampled, then 10 datasets in which 2 community members sequences were sampled, etc. up to 10 datasets in which all 57 community members sequences were sampled. For each subsampled dataset, we then combined these subsampled datasets with the background North American sequences, and reran the Nextstrain pipeline. For each subsample and trial, we infer geographic transmission history across the tree and enumerate the number of introductions into Washington. Geographic transmission history was inferred using a discrete trait model in TreeTime(Sagulenko et al., 2018). For each number of sequences tested, *n*, we report the number of trials resulting in that number of inferred introductions, and the mean number of inferred introductions across the 10 trials. Each resulting “cluster” consisted of a set of sequences that are related to one another that descend from a single inferred introduction of mumps into Washington.

### Inference of community transmission dynamics using a structured coalescent model

To infer the rates of migration between community and non-community members and to infer ancestral states of Washington internal nodes, we employed a structured coalescent model. Here, “state” refers to the inferred ancestral identity of an internal node, where the identity could be inferred as “Marshallese” or “not Marshallese”. The multitype tree model (Vaughan et al., 2014) in BEAST 2 v2.6.2 (Bouckaert et al., 2019) infers the effective population sizes of each deme and the migration rates between them. Because the multitype tree model requires that all partitions contain all demes, we could only analyze 4 clades that circulated in Washington State and included both Marshallese and non-Marshallese tips. We generated an XML in BEAUti v2.6.2 with 4 partitions, and linked the clock, site, and migration models. We used a strict, fixed clock, set to 4.17 x 10^−4^ substitutions per site year, and used an HKY substitution model with 4 gamma-distributed rate categories. This clock rate was set based on the inferred substitutions per site per year from all North American mumps genomes on nextstrain.org/mumps/na. Migration rates were inferred with the prior specified as a truncated exponential distribution with a mean of 1 and a maximum of 50. Effective population sizes were inferred with the prior specified as a truncated exponential distribution with a mean of 1, a minimum value of 0.001, and a maximum value of 10,000. All other priors were left at default values. In order to improve convergence, we employed 3 heated chains using the package CoupledMCMC (Müller and Bouckaert, 2019), where proposals for chains to swap were performed every 100 states. The analysis was run for 100 million steps, with states sampled every 1 million steps. We ran this analysis 3 independent times, and combined log and tree file output from those independent runs using LogCombiner, with the first 10% (1000 states) of each run discarded as burnin. We then summarized these combined output log and tree files. A maximum clade credibility tree was inferred using TreeAnnotator with the mean heights option. To ensure that results were not appreciably altered by the migration rate prior, we also repeated these analyses with migration rates inferred with the prior specified as a truncated exponential distribution with a mean of 10 and a maximum of 50.

Although our complete dataset contains approximately equal numbers of sequences from Marshallese and non-Marshallese cases, the 4 clusters analyzed above are enriched among Marshallese tips. To assess the impact of uneven sampling within these clusters on ancestral state inference, we performed a subsampling analysis. For each cluster, we subsampled down the number of Marshallese tips to be equal to the number of non-Marshallese tips, and reran the analysis as above. While the original analysis used 4 subclades containing both Marshallese and non-Marshallese tips, one of these subclades only has 5 tips. Subsampling this particular subtree would have resulted in a subtree with only 2 tips, thus we excluded this clade from the subsampling analysis. For this sensitivity analysis, the 3 subsampled datasets had the following tip composition: primary outbreak clade: 26 Marshallese and 26 non-Marshallese tips; 10-tip introduction: 3 Marshallese and 3 non-Marshallese tips; 8-tip introduction: 4 Marshallese and 4 non-Marshallese tips. We generated 3 randomly subsampled datasets, and for each one ran 3 independent chains, with each chain run for 50 million steps, sampling every 500,000. For one of the subsampled datasets, none of the chains converged after 20 days. In each of the remaining 2 subsampled datasets, 2 out of 3 chains converged. We combined these converged chains using LogCombiner, with the first 10% of each run discarded as burn-in. We then summarized these combined output log and tree files, and inferred a maximum clade credibility tree using TreeAnnotator with the mean heights option.

The analysis as described above assumes that each introduction into Washington State is an independent observation of the same structured coalescent process, and that the dataset represents a random sample of the underlying population. Additionally, this approach requires *a priori* definition of which sequences are part of the same Washington State transmission chain. Finally, the above analysis could only make use of the 4 Washington introductions with both Marshallese and non-Marshallese tips, and excludes other transmission chains. Because of these issues, we supplemented the above approach with an additional analysis using the approximate structured coalescent (Müller et al., 2017) in MASCOT (Müller et al., 2018). Using all of the Washington sequences, we specified three demes: Marshallese in Washington, non-Marshallese in Washington, and outside of Washington. To account for any transmission that happened outside of Washington State, the “outside of Washington” deme acted as a “ghost deme” from which we did not use any samples. The effective population size of this “outside of Washington” deme then describes the rate at which lineages between any location outside of Washington share a common ancestor. Including specific samples from outside of Washington would bias the inferred effective population size towards the coalescent rates of the sampled locations, by incorporating local transmission dynamics of other locations. We then estimated migration rates and effective population sizes for all 3 demes, but fixed the migration rates such that the unsampled deme (“outside of Washington”) could only act as a source population. This is motivated by not having observed obvious migration out of Washington State in our previous analysis here. We ran this analysis for 10 million steps, sampling every 5000, and discarded the first 10% of states as burnin.

## Acknowledgments

We would like to extend our sincerest thank you to Jiji Jally for her help and input on the project. Jiji Jally is an advocate for affordable access to healthcare services, supportive services for the Marshallese community, and works as a translator to assist the community in Washington State. These insightful discussions were absolutely critical for contextualizing our results. We would also like to sincerely thank Kelsey Florek for locating and sharing mumps samples from Wisconsin, Ohio, Missouri, Alabama, and North Carolina, which greatly enhanced the analyses presented here. We also thank Jeff Joy for graciously sharing mumps genomes from British Columbia. Finally, we would like to thank the Fred Hutchinson Cancer Research Center sequencing core for providing excellent sequencing services and Fred Hutch Scientific Computing Infrastructure. LHM is an Open Philanthropy Project fellow of the Life Sciences Research Foundation. AB was supported by the National Science Foundation Graduate Research Fellowship Program under Grant No. DGE-1256082. TB is a Pew Biomedical Scholar and is supported by NIH R35 GM119774-01. Scientific Computing Infrastructure at Fred Hutch is supported by NIH ORIP S10OD028685.

## Author contributions

LHM and AB generated and analyzed data and wrote the manuscript. CD and NRG provided data and contributed to writing the manuscript. ML, APO, and SL, provided sample and support for the study. DH provided conceptual input and contributed to the writing of the manuscript. NFM analyzed data and helped write the manuscript. TB planned the study, supported data analysis, and wrote the manuscript.

---

**Supplemental Figure 1:**
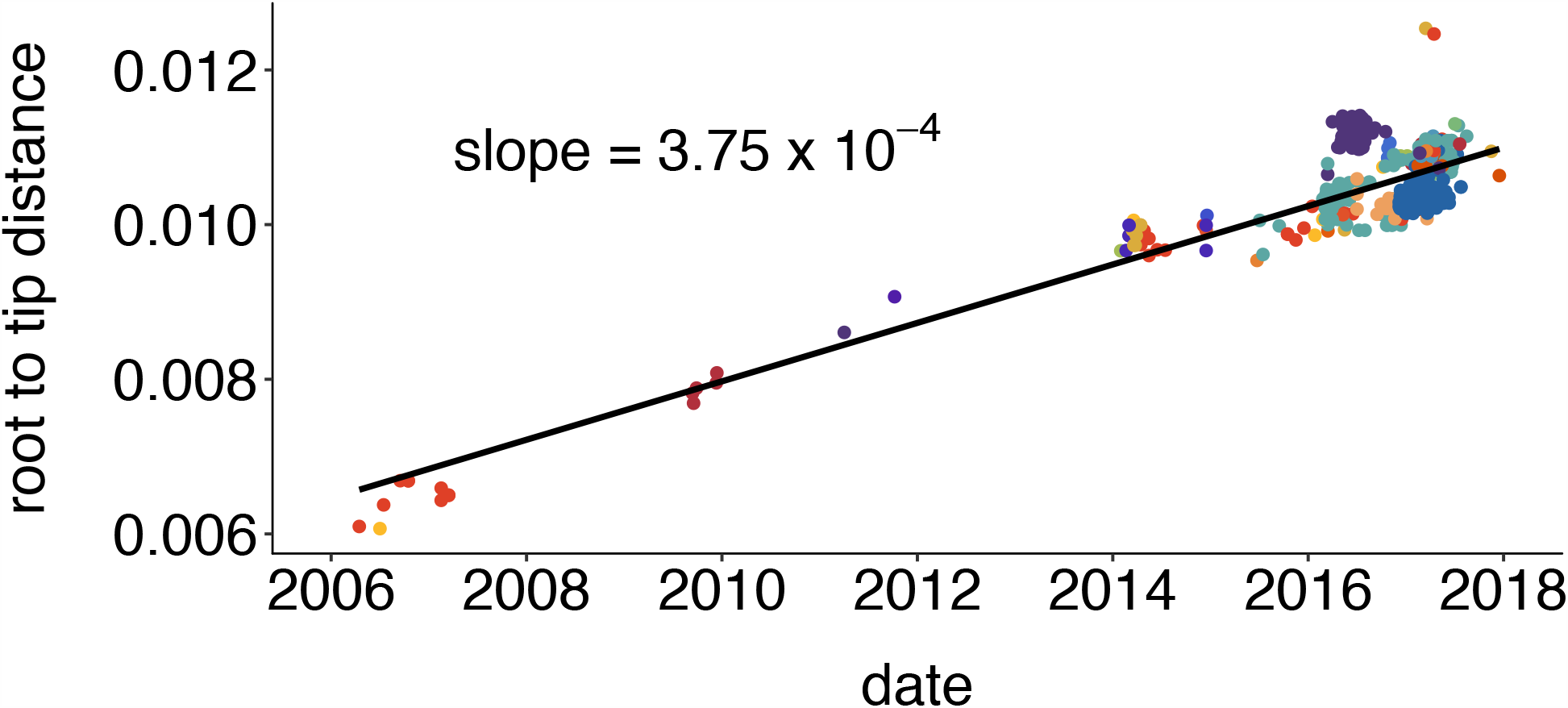
Mumps genomes accumulate mutations linearly over time. We inferred a maximum likelihood phylogeny using IQTREE for all available complete mumps genomes of genotype G, sampled from North America between 2006 and 2018. We inferred the root-to-tip distance with TempEst and plot the root to tip divergence vs. sample collection date. Color represents geographic location (either Canadian province or US state), with colors corresponding to those in Figure 2. We infer that mumps genomes accumulate mutations at a rate of 3.75 x 10^−4^ substitutions per site per year.

**Supplemental Figure 2:**
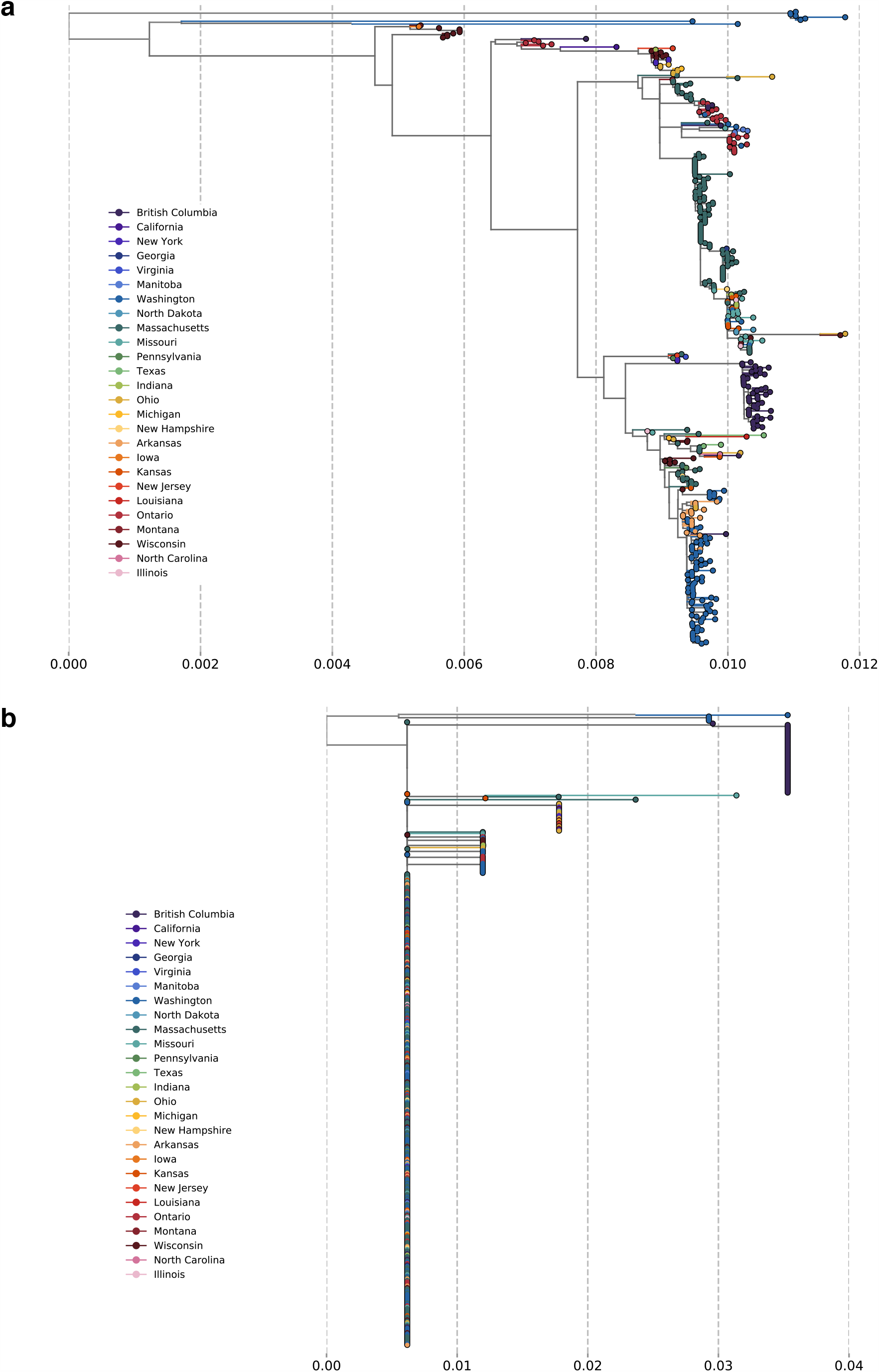
SH gene sequences are inadequate for fine-scale resolution of mumps tranmission. **a**. We inferred a maximum likelihood phylogeny using IQTREE for all available complete mumps genomes of genotype G, sampled from North America between 2006 and 2018. Color represents geographic location (either Canadian province or US state), and the x-axis displays divergence in substitutions per site per year. **b**. To compare whether similar results would have been obtained if we had only sequenced the SH gene, we truncated our sequences to include only the coding region for SH and again inferred a maximum likelihood phylogeny using the same procedure as in a. The vast majority of North American mumps sequences are identical and form a single polytomy, suggesting that SH sequencing alone provides limited resolution for inferring geographic spread.

**Supplemental Figure 3:**
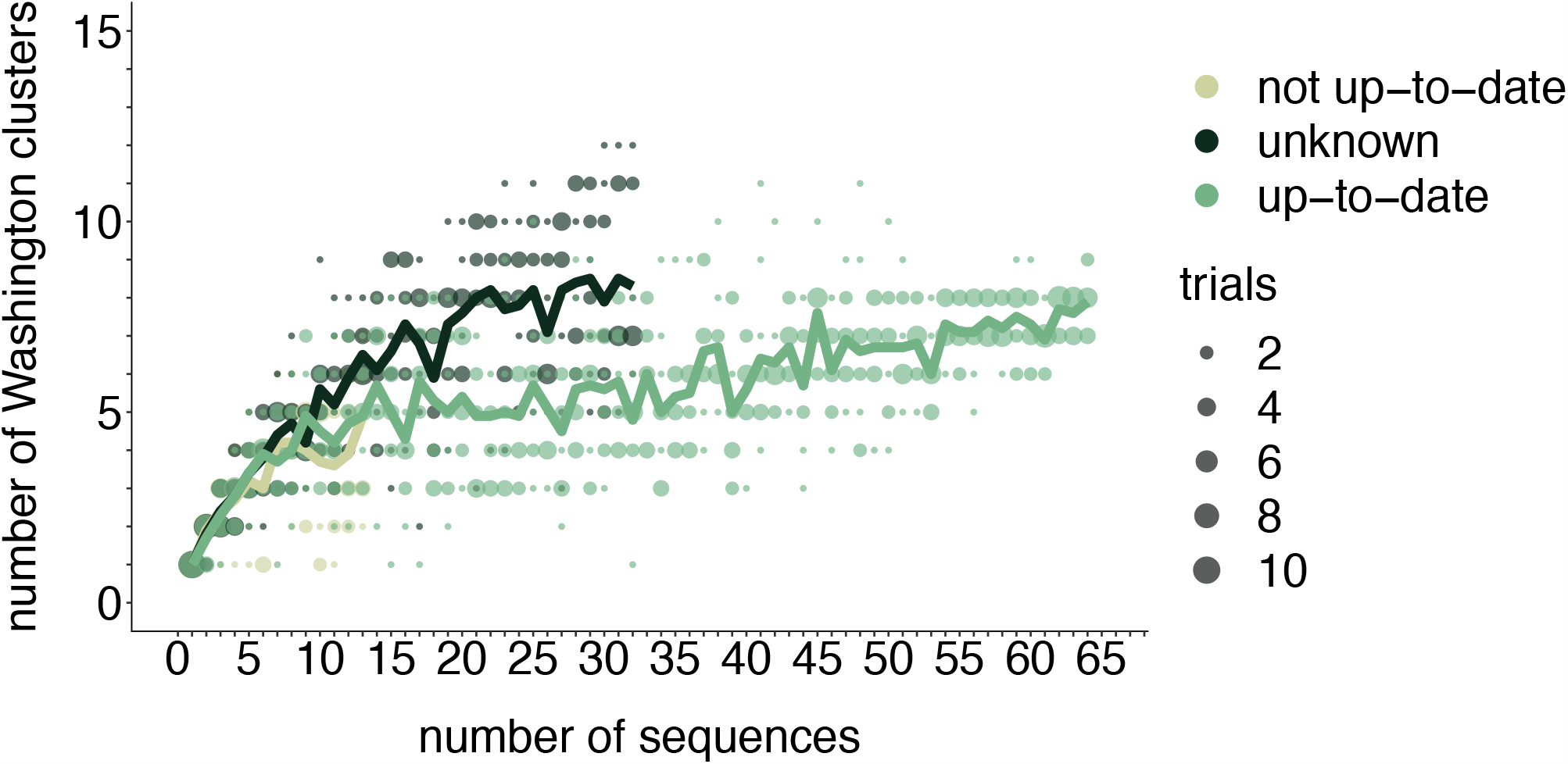
Rarefaction results by vaccination status. We repeated the rarefaction analysis shown in Figure 4b for vaccination status. We separated all Washington tips and classified them by vaccination status into up-to-date, not up-to-date, or unknown vaccination status. We then performed a rarefaction analysis and plot the number of inferred Washington clusters (y-axis) as a function of the number of sequences included in the analysis (x-axis). Dark green represents unknown vaccination status, light green represents not up-to-date, and green represents up-to-date. The majority of sequences in our dataset were derived from individuals who were up-to-date for mumps vaccine. Each dot represents the number of trials in which that number of clusters was inferred, and the solid line represents the mean across trials.

**Supplemental Figure 4:**
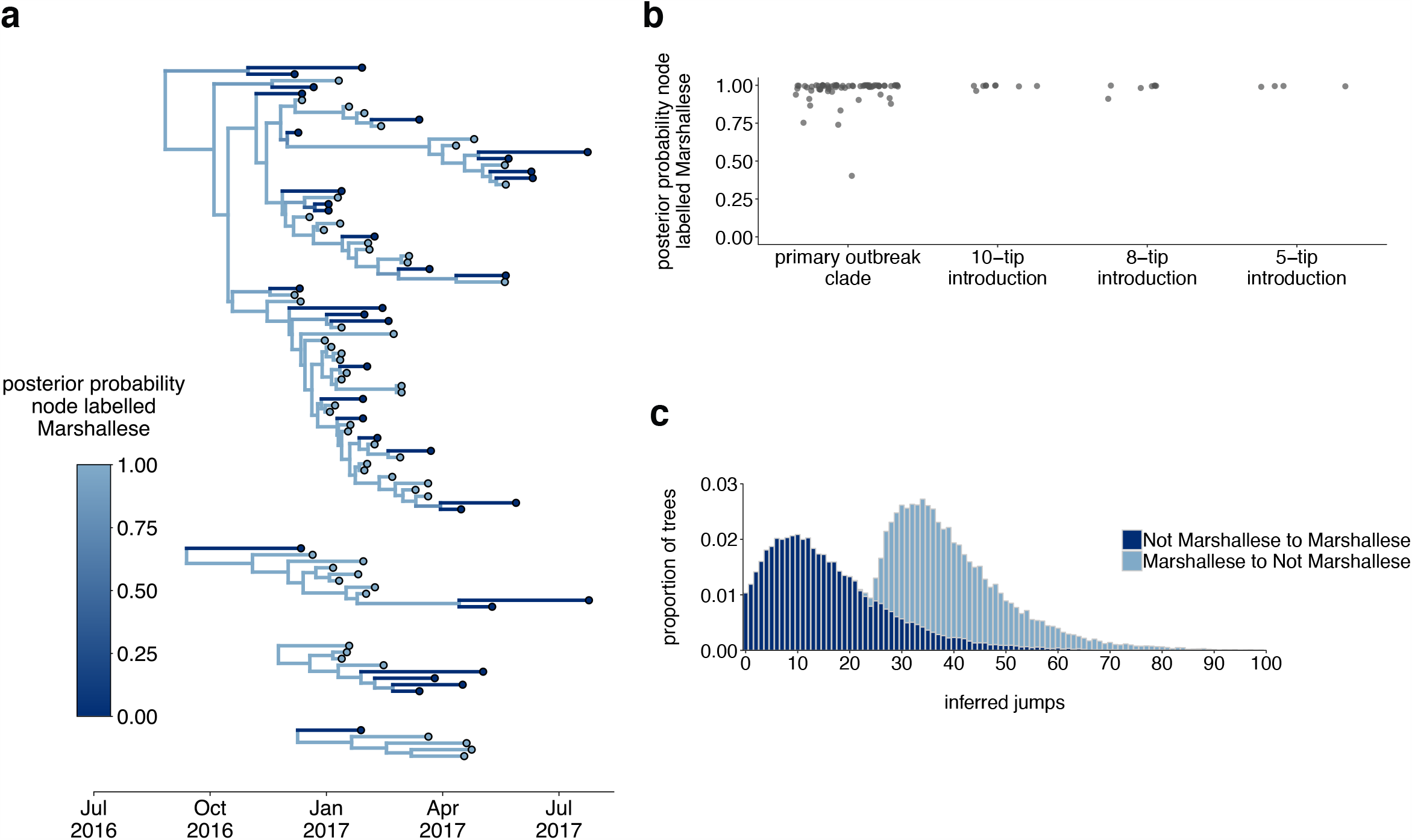
Inferences are similar under a higher migration rate prior. The results are shown for the exact same analyses displayed in Figure 5, except inferred under a model with a higher migration rate prior (10 instead of 1). **a**. Using the 4 Washington clusters that had a mixture of Marshallese and non-Marshallese cases, we inferred phylogenies using a structured coalescent model. Each group of sequences shared a clock model, migration model, and substitution model, but each topology was inferred separately, allowing us to incorporate information from all 4 clusters into the migration estimation. For each cluster, the maximum clade credibility tree is shown, where the color of each internal node represents the posterior probability that the node is Marshallese. **b**. For each internal node shown in panel **a**, we plot the posterior probability of that node being Marshallese. Across all 4 clusters, almost every internal node is inferred as Marshallese with high probability. **c**. The posterior distribution of the number of “jumps” or transmission events from Marshallese to not Marshallese (light blue) and not Marshallese to Marshallese (dark blue) inferred for the primary outbreak clade.

**Supplemental Figure 5:**
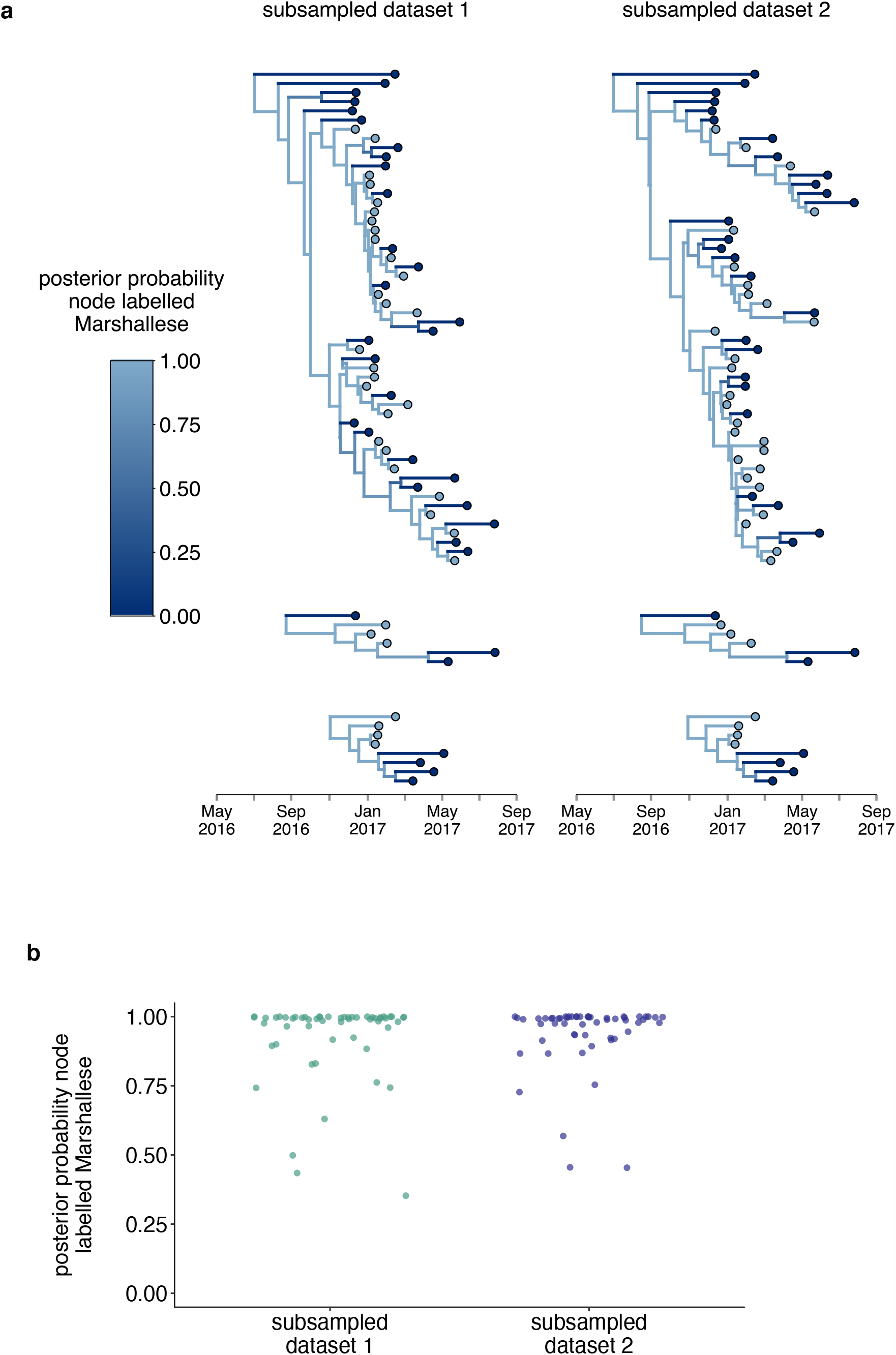
Structured coalesent analyses are robust to sampling differences. To ensure that our results were robust to differences in sampling of Marshallese and non-Marshallese tips within the clusters used for this analysis, we subsampled our dataset 3 independent times, and ran 3 independent chains per unique subsampling. In each subsampled dataset, the number of Marshallese tips was randomly subsampled to be equal to the number of non-Marshallese tips in each of the 4 clusters. We then ran each of these subsampled datasets with the exact same model as run with the full dataset. In subsampled datasets 1 and 2, 2 out of 3 chains converged, and results were combined and displayed here. In the 3rd subsampled dataset, none of the 3 chains converged, so those results are not shown. **a**. For each subsampled dataseet, we plot the inferred maximum clade credibility tree from the combined tree outputs from the 2 converged chains. The color of each tip represents whether that sample was derived from a Marshallese or non-Marshallese case, and the color of the internal node represents the posterior probability of that internal node being Marshallese. **b**. For each tree shown in **a**, the posterior probability that each internal node is labelled as Marshallese is shown. The number of the subsampled dataset is shown on the x-axis and the posterior probability is shown on the y-axis.

**Supplemental Figure 6:**
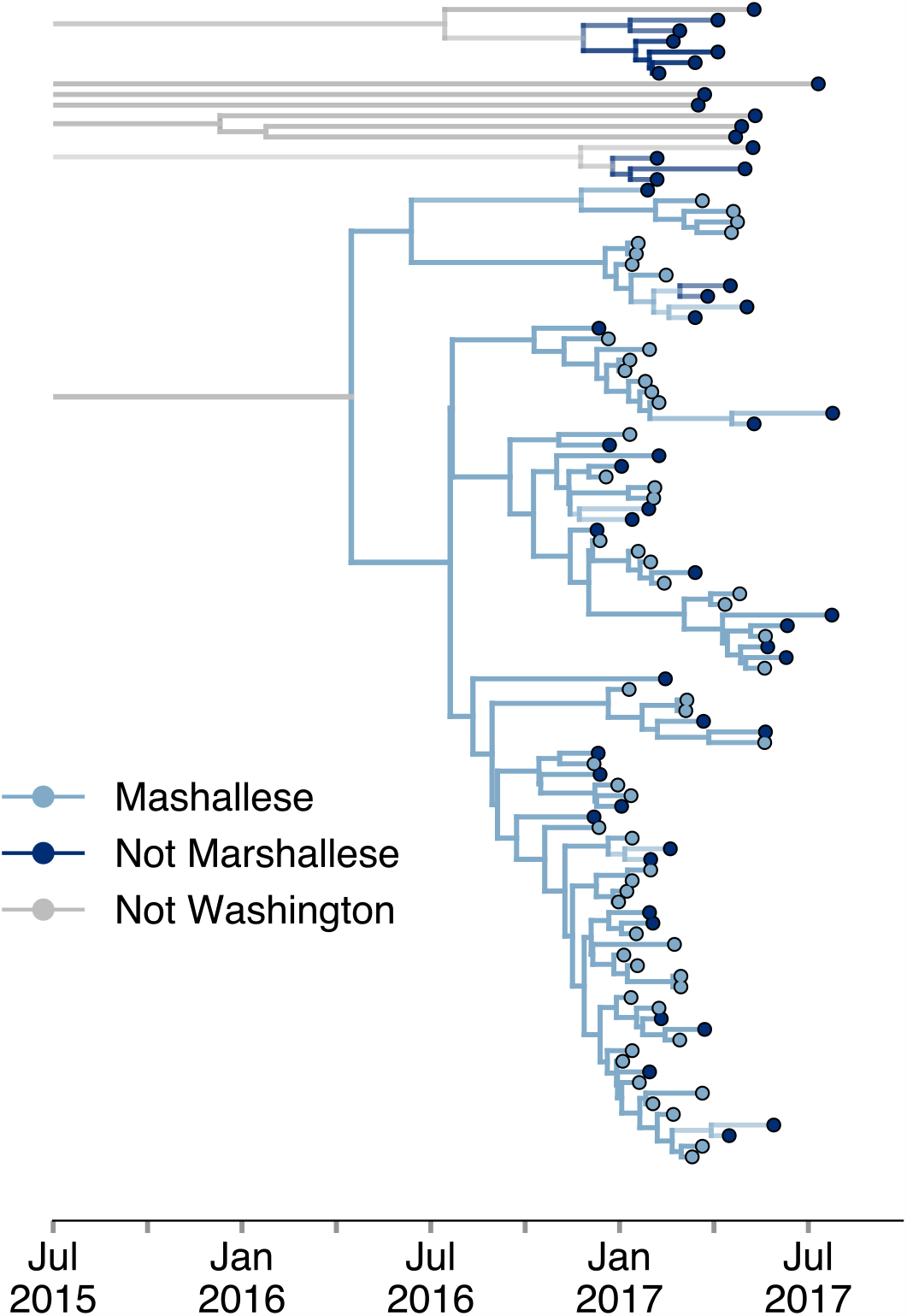
Including all Washington sequences recovers majority of transmission in Marshallese. To ensure that excluding non-Marshallese clusters did not skew our findings, we inferred a single tree using all Washington sequences. We performed a structured coalescent analysis specifying 3 groups: Marshallese, not Marshallese, and not Washington. Each internal node is colored by its most probable group, with its opacity specifying the posterior probability of being in that group (fully opaque being probability = 1, fully transparent being probability = 0).

**Supplemental Figure 7:**
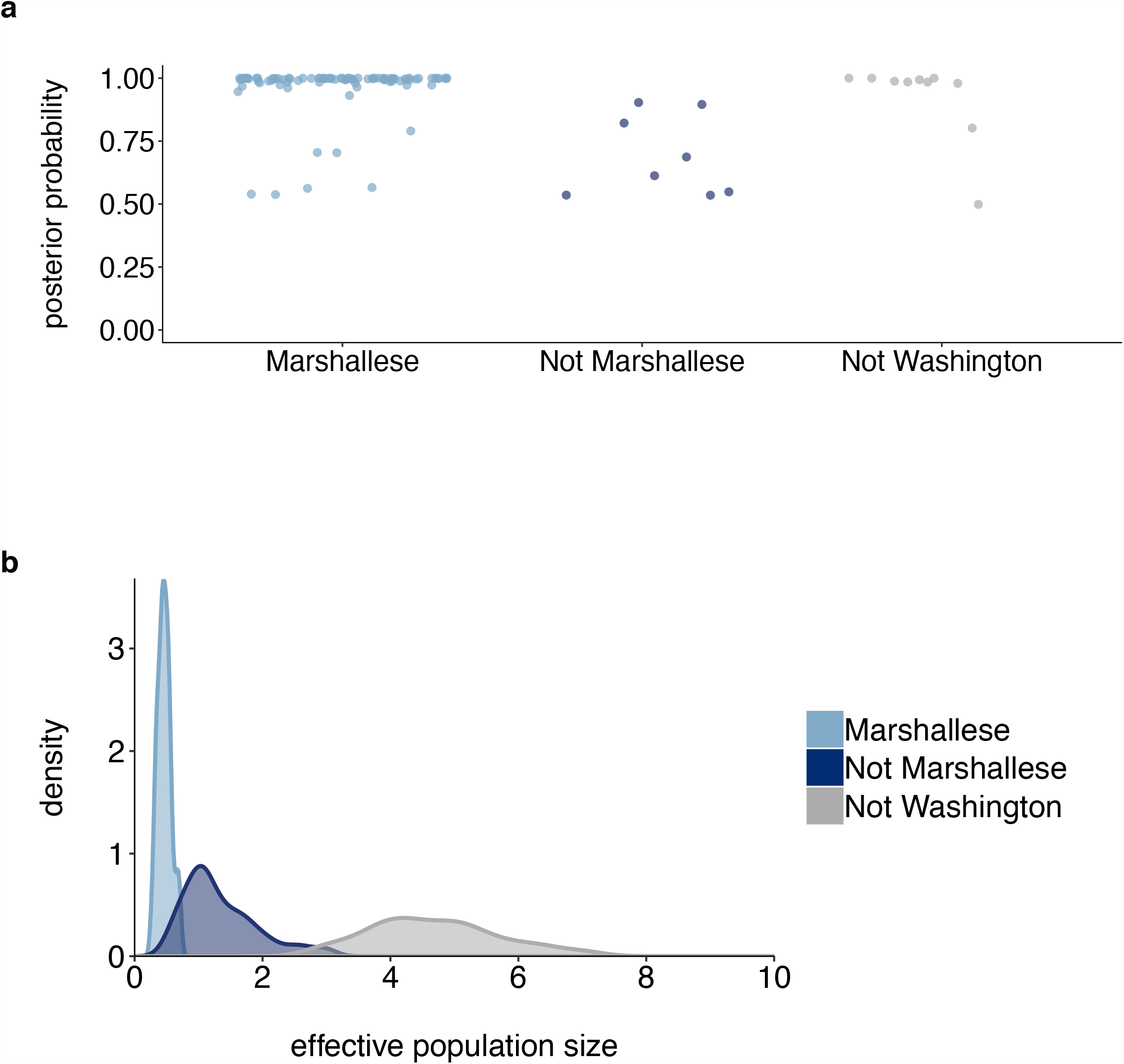
Posterior probabilities of internal node states. **a**. For the tree shown in Supplemental Figure 6, each internal node is plotted. For each intenral node, its color and placement on the x-axis represents its inferred most probable group (Marshallese, Not Marshallese, or Not Washington). The posterior probability of being labelled its most probable group is shown on the y-axis. We recover moderate support for a small number of non-Marshallese internal nodes, while the vast majority of internal nodes remain inferred as Marshallese. **b**. The 95% highest posterior density intervals of the inferred effective population sizes for Marshallese, non-Marshallese, and not Washington demes.

**Supplemental Table 1:** Sample metadata

| Strain name | Sample type | Country | US state | Mumps genotype | Vaccination status | Cycle threshold (Ct) | Genome coverage |
| --- | --- | --- | --- | --- | --- | --- | --- |
| MuVs/Washington.USA/48.16/FH1[G] | Buccal Swab | USA | Washington | G | up-to-date | 29.32 | 0.91 |
| MuVs/Washington.USA/50.16/FH2[G] | Buccal Swab | USA | Washington | G | up-to-date | 25.11 | 0.93 |
| MuVs/Washington.USA/50.16/FH3[G] | Buccal Swab | USA | Washington | G | up-to-date | 28.53 | 0.84 |
| MuVs/Washington.USA/50.16/FH4[G] | Buccal Swab | USA | Washington | G | unknown | 27.51 | 0.84 |
| MuVs/Washington.USA/1.17/FH5[G] | Buccal Swab | USA | Washington | G | up-to-date | 23.04 | 0.93 |
| MuVs/Washington.USA/1.17/FH6[G] | Buccal Swab | USA | Washington | G | not up-to-date | 32.87 | 0.93 |
| MuVs/Washington.USA/1.17/FH7[G] | Buccal Swab | USA | Washington | G | up-to-date | 25.29 | 0.92 |
| MuVs/Washington.USA/1.17/FH8[G] | Buccal Swab | USA | Washington | G | unknown | 26.61 | 0.93 |
| MuVs/Washington.USA/5.17/FH9[G] | Buccal Swab | USA | Washington | G | up-to-date | 30.38 | 0.84 |
| MuVs/Washington.USA/5.17/FH10[G] | Buccal Swab | USA | Washington | G | up-to-date | 26.72 | 0.92 |
| MuVs/Washington.USA/5.17/FH11[G] | Buccal Swab | USA | Washington | G | up-to-date | 28.17 | 0.93 |
| MuVs/Washington.USA/6.17/FH12[G] | Buccal Swab | USA | Washington | G | unknown | 25.00 | 0.93 |
| MuVs/Washington.USA/6.17/FH13[G] | Buccal Swab | USA | Washington | G | up-to-date | 25.31 | 0.93 |
| MuVs/Washington.USA/6.17/FH14[G] | Buccal Swab | USA | Washington | G | unknown | 29.04 | 0.93 |
| MuVs/Washington.USA/7.17/FH15[G] | Buccal Swab | USA | Washington | G | unknown | 33.33 | 0.92 |
| MuVs/Washington.USA/9.17/FH16[G] | Buccal Swab | USA | Washington | G | up-to-date | 23.17 | 0.93 |
| MuVs/Washington.USA/8.17/FH17[G] | Buccal Swab | USA | Washington | G | up-to-date | 28.83 | 0.92 |
| MuVs/Washington.USA/9.17/FH18[G] | Buccal Swab | USA | Washington | G | up-to-date | 31.84 | 0.84 |
| MuVs/Washington.USA/8.17/FH19[G] | Buccal Swab | USA | Washington | G | unknown | 32.16 | 0.93 |
| MuVs/Washington.USA/15.17/FH20[G] | Buccal Swab | USA | Washington | G | up-to-date | 22.25 | 0.93 |
| MuVs/Washington.USA/2.17/FH21[G] | Buccal Swab | USA | Washington | G | up-to-date | 26.50 | 0.92 |
| MuVs/Washington.USA/2.17/FH22[G] | Buccal Swab | USA | Washington | G | not up-to-date | 31.09 | 0.85 |
| MuVs/Washington.USA/2.17/FH23[G] | Buccal Swab | USA | Washington | G | up-to-date | 25.61 | 0.92 |
| MuVs/Washington.USA/3.17/FH24[G] | Buccal Swab | USA | Washington | G | up-to-date | 21.89 | 0.93 |
| MuVs/Washington.USA/3.17/FH25[G] | Buccal Swab | USA | Washington | G | up-to-date | 27.23 | 0.93 |
| MuVs/Washington.USA/2.17/FH26[G] | Buccal | USA | Washington | G | up-to-date | 29.08 | 0.93 |
|  | Swab |  |  |  |  |  |  |
| MuVs/Washington.USA/5.17/FH27[G] | Buccal Swab | USA | Washington | G | up-to-date | 24.53 | 0.93 |
| MuVs/Washington.USA/52.16/FH28[G] | Buccal Swab | USA | Washington | G | unknown | 34.28 | 0.74 |
| MuVs/Washington.USA/1.17/FH29[G] | Buccal Swab | USA | Washington | G | up-to-date | 33.14 | 0.64 |
| MuVs/Washington.USA/5.17/FH30[G] | Buccal Swab | USA | Washington | G | unknown | 33.27 | 0.63 |
| MuVs/Washington.USA/9.17/FH31[G] | Buccal Swab | USA | Washington | G | up-to-date | 34.94 | 0.66 |
| MuVs/Washington.USA/30.17/FH32[G] | Buccal Swab | USA | Washington | G | not up-to-date | 21.88 | 0.93 |
| MuVs/Washington.USA/50.16/FH33[G] | Buccal Swab | USA | Washington | G | unknown | 23.94 | 0.92 |
| MuVs/Washington.USA/12.17/FH34[G] | Buccal Swab | USA | Washington | G | up-to-date | 24.15 | 0.93 |
| MuVs/Washington.USA/12.17/FH35[G] | Buccal Swab | USA | Washington | G | not up-to-date | 28.12 | 0.93 |
| MuVs/Washington.USA/16.17/FH36[G] | Buccal Swab | USA | Washington | G | up-to-date | 25.15 | 0.93 |
| MuVs/Washington.USA/17.17/FH37[G] | Buccal Swab | USA | Washington | G | unknown | 25.71 | 0.93 |
| MuVs/Washington.USA/17.17/FH38[G] | Buccal Swab | USA | Washington | G | unknown | 26.45 | 0.93 |
| MuVs/Washington.USA/17.17/FH39[G] | Buccal Swab | USA | Washington | G | up-to-date | 26.69 | 0.93 |
| MuVs/Washington.USA/17.17/FH40[G] | Buccal Swab | USA | Washington | G | up-to-date | 21.78 | 0.93 |
| MuVs/Washington.USA/51.16/FH41[G] | Buccal Swab | USA | Washington | G | unknown | 23.56 | 0.93 |
| MuVs/Washington.USA/9.17/FH42[G] | Buccal Swab | USA | Washington | G | up-to-date | 23.29 | 0.93 |
| MuVs/Washington.USA/10.17/FH43[G] | Buccal Swab | USA | Washington | G | up-to-date | 24.50 | 1.00 |
| MuVs/Washington.USA/19.17/FH44[G] | Buccal Swab | USA | Washington | G | unknown | 27.26 | 0.93 |
| MuVs/Washington.USA/49.16/FH45[G] | Buccal Swab | USA | Washington | G | unknown | 30.24 | 0.93 |
| MuVs/Washington.USA/51.16/FH46[G] | Buccal Swab | USA | Washington | G | unknown | 25.69 | 0.93 |
| MuVs/Washington.USA/2.17/FH47[G] | Buccal Swab | USA | Washington | G | up-to-date | 24.04 | 0.93 |
| MuVs/Washington.USA/4.17/FH48[G] | Buccal Swab | USA | Washington | G | up-to-date | 20.15 | 0.93 |
| MuVs/Washington.USA/4.17/FH49[G] | Buccal Swab | USA | Washington | G | up-to-date | 31.31 | 0.93 |
| MuVs/Washington.USA/5.17/FH50[G] | Buccal Swab | USA | Washington | G | unknown | 31.14 | 0.85 |
| MuVs/Washington.USA/6.17/FH51[G] | Buccal Swab | USA | Washington | G | unknown | 26.59 | 0.92 |
| MuVs/Washington.USA/10.17/FH52[G] | Buccal Swab | USA | Washington | G | not up-to-date | 23.76 | 0.93 |
| MuVs/Washington.USA/19.17/FH53[G] | Buccal | USA | Washington | G | unknown | 22.25 | 0.93 |
| G] | Swab |  |  |  |  |  |  |
| MuVs/Washington.USA/2.17/FH54[G] | Buccal Swab | USA | Washington | G | up-to-date | 24.47 | 0.84 |
| MuVs/Washington.USA/49.16/FH55[G] | nasopharyngeal/throat swab | USA | Washington | G | unknown | 28.39 | 0.84 |
| MuVs/Washington.USA/11.17/FH56[G] | Buccal Swab | USA | Washington | G | unknown | 25.31 | 1.00 |
| MuVs/Washington.USA/12.17/FH57[G] | Buccal Swab | USA | Washington | G | unknown | 32.79 | 0.93 |
| MuVs/Washington.USA/2.17/FH58[G] | Buccal Swab | USA | Washington | G | up-to-date | 23.20 | 0.93 |
| MuVs/Washington.USA/3.17/FH59[G] | Buccal Swab | USA | Washington | G | up-to-date | 26.24 | 0.93 |
| MuVs/Washington.USA/4.17/FH60[G] | Buccal Swab | USA | Washington | G | not up-to-date | 26.87 | 0.93 |
| MuVs/Washington.USA/7.17/FH61[G] | Buccal Swab | USA | Washington | G | unknown | 24.86 | 0.98 |
| MuVs/Washington.USA/3.17/FH62[G] | Buccal Swab | USA | Washington | G | unknown | 26.99 | 0.94 |
| MuVs/Washington.USA/16.17/FH63[G] | Buccal Swab | USA | Washington | G | up-to-date | 24.26 | 0.99 |
| MuVs/Washington.USA/50.16/FH64[G] | Buccal Swab | USA | Washington | G | up-to-date | 34.54 | 0.93 |
| MuVs/Washington.USA/1.17/FH65[G] | Buccal Swab | USA | Washington | G | up-to-date | 28.53 | 0.93 |
| MuVs/Washington.USA/2.17/FH66[G] | Buccal | USA | Washington | G | unknown | 23.51 | 0.93 |
|  | Swab |  |  |  |  |  |  |
| MuVs/Washington.USA/3.17/FH67[G] | Buccal Swab | USA | Washington | G | up-to-date | 29.70 | 0.93 |
| MuVs/Washington.USA/4.17/FH68[G] | Buccal Swab | USA | Washington | G | up-to-date | 27.07 | 0.93 |
| MuVs/Washington.USA/21.17/FH69[G] | Buccal Swab | USA | Washington | G | not up-to-date | 22.46 | 0.93 |
| MuVs/Washington.USA/4.17/FH70[G] | Buccal Swab | USA | Washington | G | unknown | 25.32 | 0.93 |
| MuVs/Washington.USA/4.17/FH71[G] | Buccal Swab | USA | Washington | G | up-to-date | 25.92 | 0.93 |
| MuVs/Washington.USA/5.17/FH72[G] | Buccal Swab | USA | Washington | G | unknown | 22.74 | 0.93 |
| MuVs/Washington.USA/20.17/FH73[G] | Buccal Swab | USA | Washington | G | up-to-date | 28.59 | 0.93 |
| MuVs/Washington.USA/22.17/FH74[G] | Buccal Swab | USA | Washington | G | up-to-date | 28.84 | 0.93 |
| MuVs/Washington.USA/52.16/FH75[G] | Buccal Swab | USA | Washington | G | up-to-date | 27.02 | 0.93 |
| MuVs/Washington.USA/12.17/FH76[G] | Buccal Swab | USA | Washington | G | not up-to-date | 25.15 | 0.93 |
| MuVs/Washington.USA/1.17/FH77[G] | Buccal Swab | USA | Washington | G | up-to-date | 34.73 | 0.92 |
| MuVs/Washington.USA/12.17/FH78[G] | Buccal Swab | USA | Washington | G | unknown | 27.05 | 0.90 |
| MuVs/Washington.USA/16.17/FH79[G] | Buccal Swab | USA | Washington | G | up-to-date | 27.44 | 0.93 |
| MuVs/Washington.USA/19.17/FH80[G] | Buccal Swab | USA | Washington | G | unknown | 29.00 | 0.93 |
| MuVs/Washington.USA/20.17/FH81[G] | Buccal Swab | USA | Washington | G | not up-to-date | 27.85 | 0.93 |
| MuVs/Washington.USA/20.17/FH82[G] | Buccal Swab | USA | Washington | G | up-to-date | 33.04 | 0.84 |
| MuVs/Washington.USA/29.17/FH83[G] | Buccal Swab | USA | Washington | G | not up-to-date | 32.28 | 0.93 |
| MuVs/Washington.USA/2.17/FH84[G] | Buccal Swab | USA | Washington | G | not up-to-date | 23.08 | 0.84 |
| MuVs/Wisconsin.USA/16.14/FH85[G] | unknown | USA | Wisconsin | G | unknown | unknown | 0.76 |
| MuVs/Wisconsin.USA/20.14/FH86[G] | unknown | USA | Wisconsin | G | unknown | unknown | 0.58 |
| MuVs/Wisconsin.USA/15.06/FH87[H] | unknown | USA | Wisconsin | H | unknown | unknown | 0.53 |
| MuVs/Missouri.USA/32.16/FH88[G] | unknown | USA | Missouri | G | unknown | unknown | 0.78 |
| MuVs/Wisconsin.USA/15.17/FH89[G] | unknown | USA | Wisconsin | G | unknown | unknown | 0.78 |
| MuVs/Alabama.USA/11.17/FH90[G] | unknown | USA | Alabama | G | unknown | unknown | 0.80 |
| MuVs/Wisconsin.USA/15.06/FH91[A] | unknown | USA | Wisconsin | A | unknown | unknown | 0.71 |
| MuVs/Alabama.USA/19.17/FH92[G] | unknown | USA | Alabama | G | unknown | unknown | 0.77 |
| MuVs/Wisconsin.USA/37.06/FH93[G] | unknown | USA | Wisconsin | G | unknown | unknown | 0.69 |
| MuVs/Washington.USA/9.17/FH94[K] | Buccal Swab | USA | Washington | K | unknown | 29.13 | 0.63 |
| MuVs/Washington.USA/11.17/FH95[G] | Buccal Swab | USA | Washington | G | up-to-date | 28.52 | 0.77 |
| MuVs/Washington.USA/11.17/FH96[G] | Buccal Swab | USA | Washington | G | up-to-date | 36.91 | 0.76 |
| MuVs/Washington.USA/12.17/FH97[G] | Buccal Swab | USA | Washington | G | up-to-date | 35.13 | 0.63 |
| MuVs/Washington.USA/14.17/FH98[G] | Buccal Swab | USA | Washington | G | not up-to-date | 29.59 | 0.76 |
| MuVs/Wisconsin.USA/41.06/FH99[G] | unknown | USA | Wisconsin | G | unknown | unknown | 0.77 |
| MuVs/Washington.USA/19.17/FH100[G] | Buccal Swab | USA | Washington | G | up-to-date | 29.44 | 0.78 |
| MuVs/Missouri.USA/28.17/FH101[G] | unknown | USA | Missouri | G | unknown | unknown | 0.80 |
| MuVs/Ohio.USA/2.18/FH102[G] | unknown | USA | Ohio | G | unknown | unknown | 0.76 |
| MuVs/Washington.USA/4.17/FH103[G] | Buccal Swab | USA | Washington | G | up-to-date | 34.19 | 0.56 |
| MuVs/Washington.USA/5.17/FH104[G] | Buccal Swab | USA | Washington | G | up-to-date | 31.96 | 0.74 |
| MuVs/Washington.USA/6.17/FH105[G] | Buccal Swab | USA | Washington | G | up-to-date | 36.53 | 0.65 |
| MuVs/Washington.USA/7.17/FH106[G] | Buccal Swab | USA | Washington | G | not up-to-date | 27.98 | 0.67 |
| MuVs/Wisconsin.USA/7.07/FH107[G] | unknown | USA | Wisconsin | G | unknown | unknown | 0.80 |
| MuVs/Wisconsin.USA/11.07/FH108[G] | unknown | USA | Wisconsin | G | unknown | unknown | 0.94 |
| MuVs/Wisconsin.USA/11.07/FH109[G] | unknown | USA | Wisconsin | G | unknown | unknown | 0.94 |
| MuVs/Wisconsin.USA/16.14/FH110[G] | unknown | USA | Wisconsin | G | unknown | unknown | 0.92 |
| MuVs/Wisconsin.USA/20.14/FH111[G] | unknown | USA | Wisconsin | G | unknown | unknown | 0.94 |
| MuVs/Wisconsin.USA/24.14/FH112[G] | unknown | USA | Wisconsin | G | unknown | unknown | 0.87 |
| MuVs/Wisconsin.USA/29.14/FH113[G] | unknown | USA | Wisconsin | G | unknown | unknown | 0.93 |
| MuVs/Missouri.USA/29.15/FH114[G] | unknown | USA | Missouri | G | unknown | unknown | 0.84 |
| MuVs/Wisconsin.USA/42.15/FH115[G] | unknown | USA | Wisconsin | G | unknown | unknown | 0.87 |
| MuVs/Wisconsin.USA/42.15/FH116[G] | unknown | USA | Wisconsin | G | unknown | unknown | 0.88 |
| MuVs/Wisconsin.USA/46.15/FH117[G] | unknown | USA | Wisconsin | G | unknown | unknown | 0.84 |
| MuVs/Wisconsin.USA/2.16/FH118[G] | unknown | USA | Wisconsin | G | unknown | unknown | 0.91 |
| MuVs/Ohio.USA/19.16/FH119[G] | unknown | USA | Ohio | G | unknown | unknown | 0.93 |
| MuVs/Wisconsin.USA/24.16/FH120[G] | unknown | USA | Wisconsin | G | unknown | unknown | 0.89 |
| MuVs/Wisconsin.USA/19.16/FH121[G] | unknown | USA | Wisconsin | G | unknown | unknown | 0.93 |
| MuVs/Missouri.USA/41.16/FH122[G] | unknown | USA | Missouri | G | unknown | unknown | 0.85 |
| MuVs/Missouri.USA/46.16/FH123[G] | unknown | USA | Missouri | G | unknown | unknown | 0.88 |
| MuVs/Missouri.USA/46.16/FH124[G] | unknown | USA | Missouri | G | unknown | unknown | 0.90 |
| MuVs/Missouri.USA/50.16/FH125[G] | unknown | USA | Missouri | G | unknown | unknown | 0.88 |
| MuVs/Wisconsin.USA/50.16/FH126[G] | unknown | USA | Wisconsin | G | unknown | unknown | 0.88 |
| MuVs/Missouri.USA/2.17/FH127[G] | unknown | USA | Missouri | G | unknown | unknown | 0.83 |
| MuVs/Ohio.USA/2.17/FH128[G] | unknown | USA | Ohio | G | unknown | unknown | 0.90 |
| MuVs/Missouri.USA/7.17/FH129[G] | unknown | USA | Missouri | G | unknown | unknown | 0.84 |
| MuVs/Ohio.USA/7.17/FH130[G] | unknown | USA | Ohio | G | unknown | unknown | 0.89 |
| MuVs/Wisconsin.USA/15.06/FH131[G] | unknown | USA | Wisconsin | G | unknown | unknown | 0.81 |
| MuVs/Ohio.USA/11.17/FH132[G] | unknown | USA | Ohio | G | unknown | unknown | 0.86 |
| MuVs/Missouri.USA/11.17/FH133[G] | unknown | USA | Missouri | G | unknown | unknown | 0.85 |
| MuVs/Missouri.USA/15.17/FH134[G] | unknown | USA | Missouri | G | unknown | unknown | 0.85 |
| MuVs/Missouri.USA/15.17/FH135[G] | unknown | USA | Missouri | G | unknown | unknown | 0.86 |
| MuVs/NorthCarolina.USA/11.17/FH136[G] | unknown | USA | NorthCarolina | G | unknown | unknown | 0.84 |
| MuVs/Wisconsin.USA/15.17/FH137[G] | unknown | USA | Wisconsin | G | unknown | unknown | 0.85 |
| MuVs/Missouri.USA/15.17/FH138[G] | unknown | USA | Missouri | G | unknown | unknown | 0.82 |
| MuVs/Alabama.USA/15.17/FH139[G] | unknown | USA | Alabama | G | unknown | unknown | 0.83 |
| MuVs/Missouri.USA/19.17/FH140[G] | unknown | USA | Missouri | G | unknown | unknown | 0.84 |
| MuVs/Wisconsin.USA/19.17/FH141[G] | unknown | USA | Wisconsin | G | unknown | unknown | 0.85 |
| MuVs/Washington.USA/8.17/FH142[G] | Buccal Swab | USA | Washington | G | up-to-date | 25.33 | 0.86 |
| MuVs/Washington.USA/11.17/FH143[G] | Buccal Swab | USA | Washington | G | up-to-date | 28.34 | 0.87 |
| MuVs/Washington.USA/12.17/FH144[G] | Buccal Swab | USA | Washington | G | unknown | 29.66 | 0.85 |
| MuVs/Washington.USA/14.17/FH145[G] | Buccal Swab | USA | Washington | G | up-to-date | 21.45 | 0.91 |
| MuVs/Washington.USA/15.17/FH146[G] | Buccal Swab | USA | Washington | G | up-to-date | 27.09 | 0.84 |
| MuVs/Washington.USA/18.17/FH147[G] | Buccal Swab | USA | Washington | G | up-to-date | 28.86 | 0.88 |
| MuVs/Washington.USA/20.17/FH148[G] | Buccal Swab | USA | Washington | G | up-to-date | 26.59 | 0.82 |
| MuVs/Washington.USA/23.17/FH149[G] | Buccal Swab | USA | Washington | G | unknown | 33.05 | 0.84 |
| MuVs/Washington.USA/23.17/FH150[G] | Buccal Swab | USA | Washington | G | up-to-date | 32.33 | 0.84 |
| MuVs/Washington.USA/28.17/FH151[G] | Buccal Swab | USA | Washington | G | up-to-date | 24.18 | 0.88 |
| MuVs/Washington.USA/16.17/FH152[G] | Buccal Swab | USA | Washington | G | up-to-date | 27.41 | 0.85 |
| MuVs/Wisconsin.USA/41.06/FH153[G] | unknown | USA | Wisconsin | G | unknown | unknown | 0.84 |
| MuVs/Wisconsin.USA/7.07/FH154[G] | unknown | USA | Wisconsin | G | unknown | unknown | 0.94 |
| MuVs/Missouri.USA/33.17/FH155[G] | unknown | USA | Missouri | G | unknown | unknown | 0.85 |
| MuVs/Alabama.USA/50.17/FH156[G] | unknown | USA | Alabama | G | unknown | unknown | 0.85 |
| MuVs/Ohio.USA/46.17/FH157[G] | unknown | USA | Ohio | G | unknown | unknown | 0.84 |
| MuVs/Washington.USA/49.16/FH158[G] | Buccal Swab | USA | Washington | G | unknown | 30.55 | 0.84 |
| MuVs/Wisconsin.USA/28.06/FH159[G] | unknown | USA | Wisconsin | G | unknown | unknown | 0.84 |
| MuVs/Washington.USA/1.17/FH160[G] | Buccal Swab | USA | Washington | G | unknown | 24.17 | 0.87 |
| MuVs/Washington.USA/5.17/FH161[G] | Buccal Swab | USA | Washington | G | up-to-date | 23.05 | 0.92 |
| MuVs/Washington.USA/5.17/FH162[G] | Buccal Swab | USA | Washington | G | up-to-date | 27.08 | 0.90 |
| MuVs/Wisconsin.USA/51.15/FH163[G] | unknown | USA | Wisconsin | G | unknown | unknown | 0.86 |
| MuVs/Alabama.USA/7.17/FH164[G] | unknown | USA | Alabama | G | unknown | unknown | 0.80 |
| MuVs/Missouri.USA/24.17/FH165[G] | unknown | USA | Missouri | G | unknown | unknown | 0.86 |
| MuVs/Washington.USA/7.17/FH166[G] | Buccal Swab | USA | Washington | G | up-to-date | 27.96 | 0.85 |

All genomes generated for this analysis are described above. Dates are formatted as Year-month-day. Vaccination status, Ct, and sample collection type are all available for the Washington samples. Genome coverage represents the total proportion of bases in the genome with at least 20x coverage for which we were able to call a base. Sites with <20x coverage were labelled as Ns. Only samples with at least 50% non-N bases were included in the analysis.

**Supplemental Table 2:** Mumps cases by age group.

| Age group | Mumps case count | 2016-2017 average population in Washington<sup>1</sup> | Rate of cases per 100,000 individuals | Percentage of total cases |
| --- | --- | --- | --- | --- |
| 0 - 4 | 34 | 450,847 | 7.5 | 3.8% |
| 5 - 9 | 104 | 462,951 | 22.5 | 11.7% |
| 10 - 14 | 198 | 451,485 | 43.9 | 22.3% |
| 15 - 19 | 214 | 455,612 | 47.0 | 24.1% |
| 20 - 39 | 256 | 1,980,004 | 12.9 | 28.8% |
| 40 - 64 | 80 | 2,348,529.5 | 3.4 | 10.0% |
| 65+ | 2 | 1,097,571 | 0.2 | 0.22% |
| Unknown | 1 | NA | NA | 0.11% |

**Supplemental Table 3:** Outbreak characteristic and dataset composition.

|  | counts in outbreak (%) | Count in dataset (%) |
| --- | --- | --- |
| Up-to-date vaccination | 574 (65%) | 64 (58%) |
| Not up-to-date vaccination | 86 (9.7%) | 13 (12%) |
| Unknown vaccination status | 229 (26%) | 33 (30%) |
| Marshallese | 465 (52%) | 57 (52%) |
| Not Marshallese | 424 (48%) | 53 (48%) |
| **Total** | **889** | **110** |

